# Interpretation of temporal and spatial trends of SARS-CoV-2 RNA in San Francisco Bay Area wastewater

**DOI:** 10.1101/2021.05.04.21256418

**Authors:** Hannah D. Greenwald, Lauren C. Kennedy, Adrian Hinkle, Oscar N. Whitney, Vinson B. Fan, Alexander Crits-Christoph, Sasha Harris-Lovett, Avi I. Flamholz, Basem Al-Shayeb, Lauren D. Liao, Matt Beyers, Daniel Brown, Alicia R. Chakrabarti, Jason Dow, Dan Frost, Mark Koekemoer, Chris Lynch, Payal Sarkar, Eileen White, Rose Kantor, Kara L. Nelson

## Abstract

Wastewater surveillance for severe acute respiratory syndrome coronavirus 2 (SARS-CoV-2) RNA can be integrated with COVID-19 case data to inform timely pandemic response. However, more research is needed to apply and develop systematic methods to interpret the true SARS-CoV-2 signal from noise introduced in wastewater samples (e.g., from sewer conditions, sampling and extraction methods, etc.). In this study, raw wastewater was collected weekly from five sewersheds and one residential facility, and wastewater SARS-CoV-2 concentrations were compared to geocoded COVID-19 clinical testing data. SARS-CoV-2 was reliably detected (95% positivity) in frozen wastewater samples when reported daily new COVID-19 cases were 2.4 or more per 100,000 people. To adjust for variation in sample fecal content, crAssphage, pepper mild mottle virus, *Bacteroides* ribosomal RNA (rRNA), and human 18S rRNA were evaluated as normalization biomarkers, and crAssphage displayed the least spatial and temporal variability. Both unnormalized SARS-CoV-2 RNA signal and signal normalized to crAssphage had positive and significant correlation with clinical testing data (Kendall’s Tau-b (*τ*)=0.43 and 0.38, respectively). Locational dependencies and the date associated with testing data impacted the lead time of wastewater for clinical trends, and no lead time was observed when the sample collection date (versus the result date) was used for both wastewater and clinical testing data. This study supports that trends in wastewater surveillance data reflect trends in COVID-19 disease occurrence and presents approaches that could be applied to make wastewater signal more interpretable and comparable across studies.

## 1. INTRODUCTION

Increasing hospitalizations and limited diagnostic testing capacity early in the coronavirus disease 2019 (COVID-19) pandemic made it clear that multiple methods to monitor circulation of severe acute respiratory syndrome coronavirus 2 (SARS-CoV-2) are needed (Bivins et al., 2020). One such method is wastewater-based epidemiology (WBE), which has provided community-scale information on drug use, personal care products, antibiotic resistance, and pathogen circulation (Choi et al., 2018). SARS-CoV-2 is a promising candidate for WBE because its RNA is detected in stool of infected individuals (Li et al., 2021; Parasa et al., 2020), and wastewater surveillance has been shown to provide early detection of population-level increases in occurrence compared to clinical data in some locations (Ahmed et al., 2021; D’Aoust et al., 2021a; Gerrity et al., 2021; Hata and Honda, 2020; Medema et al., 2020; Nemudryi et al., 2020; Peccia et al., 2020; Randazzo et al., 2020b, 2020a).

Together, wastewater and clinical testing might provide more reliable information about disease burden in communities than either method alone. Clinical testing of individuals is resource-intensive and has well-known biases (e.g., selection bias based on symptom severity, symptom recognition, occupation, etc.) (Catalogue of Bias Collaboration et al., 2017; Griffith et al., 2020; Sims and Kasprzyk-Hordern, 2020), which have compounded negative impacts in communities with higher proportions of low-income residents and of Black, Indigenous, and People of Color, including in the San Francisco Bay Area (Chamie et al., 2020; Misa et al., 2020). In contrast, WBE may provide a less biased assessment of COVID-19 occurrence (Murakami et al., 2020; Sims and Kasprzyk-Hordern, 2020). For COVID-19 WBE to be useful for public health decision-making, a better understanding is needed of the variability of SARS-CoV-2 in wastewater and how it relates to the true incidence or prevalence of COVID-19 in the contributing population (McClary-Gutierrez et al., 2021). Sources of target signal variability in wastewater include inconsistencies in sample collection and laboratory processing (Ahmed et al., 2020d; Feng et al., 2021), nucleic acid degradation based on travel time and conditions in the sewer (Hart and Halden, 2020a), and signal dilution due to rainfall and diurnal flow changes (Zahedi et al., 2021). Researchers have addressed some of these sources of variability through normalization to biomarkers, increased sampling frequency, processing biological replicates, and smoothing/forecasting (D’Aoust et al., 2021b; Feng et al., 2021; Graham et al., 2020; McLellan et al., 2021; Nemudryi et al., 2020; Stadler et al., 2020).

Normalization of target signal to flow, population, and/or an endogenous biomarker has the potential to reduce variability and scale values for comparisons across samples and locations. Across WBE studies, researchers have normalized wastewater concentrations to flow rate and population to calculate a per capita load (Chen et al., 2014; Choi et al., 2018; Zuccato et al., 2005; Zuccato Ettore et al., 2008) or to a chemical parameter (e.g., caffeine) (Been et al., 2014; Choi et al., 2018; D’Aoust et al., 2021b; Polo et al., 2020). More recently, four biological markers have emerged as promising candidates to normalize SARS-CoV-2 RNA signal for fecal content. Pepper mild mottle virus (PMMoV), a nonenveloped RNA plant virus, is commonly used for COVID-19 WBE (D’Aoust et al., 2021b; Feng et al., 2021; Whitney et al., 2021; Wu et al., 2020) but concentrations in sewage vary with season and local diet (Symonds et al., 2019). Another normalization biomarker is the cross-assembly phage (crAssphage), a non-enveloped, DNA virus that ubiquitously infects the human gut commensal bacteria *Bacteroides* (Edwards et al., 2019; Green et al., 2020; Stachler et al., 2017; Wilder et al., 2021). In addition, *Bacteroides* HF183 16S rRNA gene is widely used for detecting fecal contamination in environmental waters (Green et al., 2020; Shanks et al., 2008), and recent studies (D’Aoust et al., 2021b; Kapoor et al., 2015; Pitkänen et al., 2013) have targeted HF183 rRNA (versus the rRNA gene) to increase the sensitivity of the assay (D’Aoust et al., 2021b; Feng et al., 2021). Lastly, the human 18S ribosomal subunit RNA (18S rRNA) assay has been proposed as a normalization biomarker because it targets human cells that are shed in feces (D’Aoust et al., 2021b; Whitney et al., 2021). While each of these normalization biomarkers has been assessed independently, they have not all been compared within the same study.

In addition to normalizing the target signal, smoothing procedures can assist in discerning temporal trends in SARS-CoV-2 occurrence. While seven-day moving averages have been widely used for assessing clinical data trends in real-time (“Track Testing Trends,” n.d.), wastewater sampling is often performed only 1-3 times per week. Therefore, smoothing techniques are needed that can be applied to data with lower sampling frequency that minimize loss of temporal resolution, such as locally weighted scatterplot smoothing (Lowess) (Gibas et al., 2021; Gonzalez et al., 2020; Nemudryi et al., 2020; Vallejo et al., 2020). However, no standard value for the bandwidth parameter exists (analogous to the selection of a seven-day window for moving averages of clinical data), and the default parameter value differs between two common languages used for data analysis (R (“Source code for spatialEco package,” n.d.): 0.75 and Python (“Source code for statsmodels module,” n.d.): 0.67). Furthermore, the bandwidth selection process generally has not been specified in studies incorporating Lowess (Gibas et al., 2021; Gonzalez et al., 2020; Nemudryi et al., 2020; Vallejo et al., 2020; Wu et al., 2020).

Systematic approaches are also needed to estimate the minimum number of clinical COVID-19 cases for which SARS-CoV-2 RNA is reliably detected in wastewater (WBE case detection limit). The WBE case detection limit is dependent on the methods used to extract genetic material as well as the extent of local clinical testing and may require sewershed-specific assessment. However, a systematic approach to estimate this value across studies can aid interpretation of nondetects and elucidate the number of COVID-19 cases per capita above which COVID-19 WBE will be a reliable public health surveillance strategy. In a recent study (Wu et al., 2021), a WBE case detection limit was estimated using a dataset with 1,687 samples, which was large enough to include repeated wastewater measurements at low case numbers. With fewer data points, researchers have estimated this value observationally by reporting the number of cases they were able to detect or quantify (Hata and Honda, 2020; Medema et al., 2020).

The goal of this research was to develop and assess approaches for COVID-19 WBE data validation and interpretation. Specific objectives were to: (i) evaluate normalization biomarkers (crAssphage, pepper mild mottle virus, Bacteroides rRNA, and human 18S rRNA) for adjusting SARS-CoV-2 RNA signal to account for variable wastewater fecal content; (ii) assess SARS-CoV-2 wastewater testing as a complement to clinical testing for public health surveillance by determining the correlation between these two methods; (iii) determine whether wastewater trends lead clinical trends and could provide early warning of COVID-19 outbreaks; (iv) evaluate a systematic method for trendline smoothing; (v) develop a systematic method for estimating a WBE case detection limit; and (vi) apply these methods to interpret spatial and temporal trends in COVID-19 occurrence based on wastewater and clinical testing data. We analyzed a sample set from six locations in the San Francisco Bay Area containing 5 months of weekly raw wastewater samples paired with geocoded clinical data.

## 2. MATERIALS AND METHODS

Six locations in the San Francisco Bay area were sampled (referred to throughout as locations A, S, N, K, Q, and E). Raw wastewater was collected weekly and archived from April to September 2020, and biological replicates were processed for some locations as indicated in **Table 1**. SARS-CoV-2 and normalization biomarkers (crAssphage, PMMoV, Bacteroides rRNA, and 18S rRNA) were measured in wastewater samples via RT-qPCR. Associated physicochemical data were collected by wastewater utilities, and associated geocoded clinical COVID-19 data were collected by public health departments (Table 1).

**Table 1:**
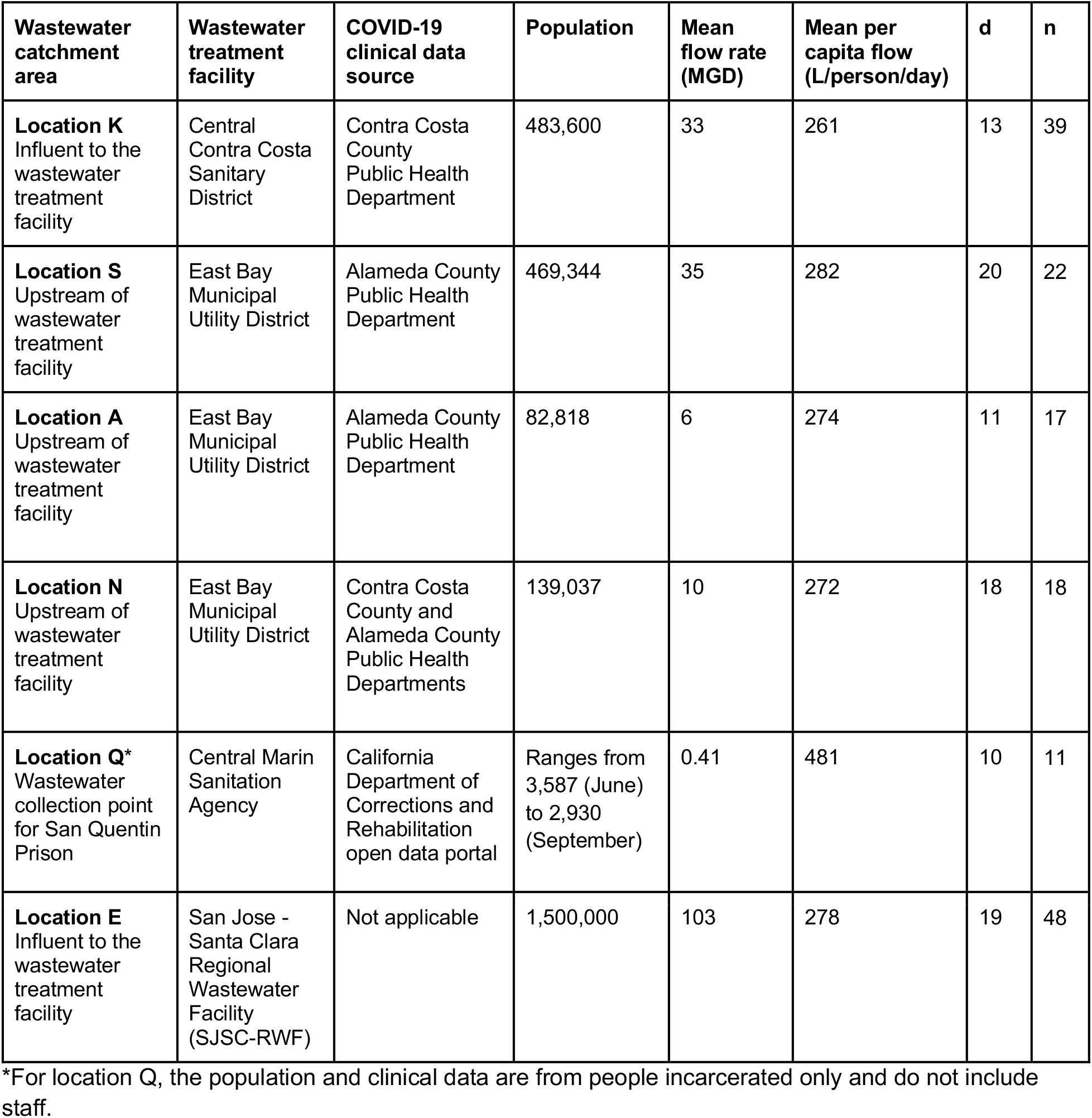
Descriptions of wastewater sampling locations including associated wastewater utility, clinical testing data sources, population, and flow rates. “d” represents the number of unique dates on which samples were collected. “n” represents the total number of wastewater samples collected, including biological replicates.

| Wastewater catchment area | Wastewater treatment facility | COVID-19 clinical data source | Population | Mean flow rate (MGD) | Mean per capita flow (L/person/day) | d | n |
| --- | --- | --- | --- | --- | --- | --- | --- |
| **Location K**<br>Influent to the wastewater treatment facility | Central Contra Costa Sanitary District | Contra Costa County Public Health Department | 483,600 | 33 | 261 | 13 | 39 |
| **Location S**<br>Upstream of wastewater treatment facility | East Bay Municipal Utility District | Alameda County Public Health Department | 469,344 | 35 | 282 | 20 | 22 |
| **Location A**<br>Upstream of wastewater treatment facility | East Bay Municipal Utility District | Alameda County Public Health Department | 82,818 | 6 | 274 | 11 | 17 |
| **Location N**<br>Upstream of wastewater treatment facility | East Bay Municipal Utility District | Contra Costa County and Alameda County Public Health Departments | 139,037 | 10 | 272 | 18 | 18 |
| **Location Q\***<br>Wastewater collection point for San Quentin Prison | Central Marin Sanitation Agency | California Department of Corrections and Rehabilitation open data portal | Ranges from 3,587 (June) to 2,930 (September) | 0.41 | 481 | 10 | 11 |
| **Location E**<br>Influent to the wastewater treatment facility | San Jose - Santa Clara Regional Wastewater Facility (SJSC-RWF) | Not applicable | 1,500,000 | 103 | 278 | 19 | 48 |

\*For location Q, the population and clinical data are from people incarcerated only and do not include staff.

### 2.1 Wastewater sample collection and physicochemical data

24-hour time-weighted composite samples of raw wastewater were collected using Teledyne ISCO autosamplers. Some samples were collected and processed in biological replicate (i.e., wastewater subsamples were aliquoted from the same composite sample but independently extracted). After collection, all samples were transported to the lab on ice, stored at either -20°C or -80°C, and then thawed at 4°C for 36-48 hours before extractions. Wastewater data was not individually identifiable; therefore, no IRB was needed. More information on location-specific data collection and wastewater sampling, transport, storage, and biological replicates is provided in **Table 1** and the **Supplementary Information (SI) Section A**.

One rainfall event occurred (May 12-19) during which sampling locations experienced 0.8 to 1.8 inches of precipitation (<u>NOAA Climate Data Online database</u>). Although none of the sampled locations was a combined sewer system, rainfall could still increase flow rates through infiltration and inflow. Daily wastewater flow rate values during this period varied <4% (Locations K & E, **Table S1**), which is negligible when compared to the variation displayed by normalization biomarkers over time (15%-244%; **Figure 1**). Mean flow rates were provided by the wastewater utility for locations A, N, and S and were calculated from daily flow rates for locations K, E, and Q (**Table 1** and **SI Section A**).

**Figure 1:**
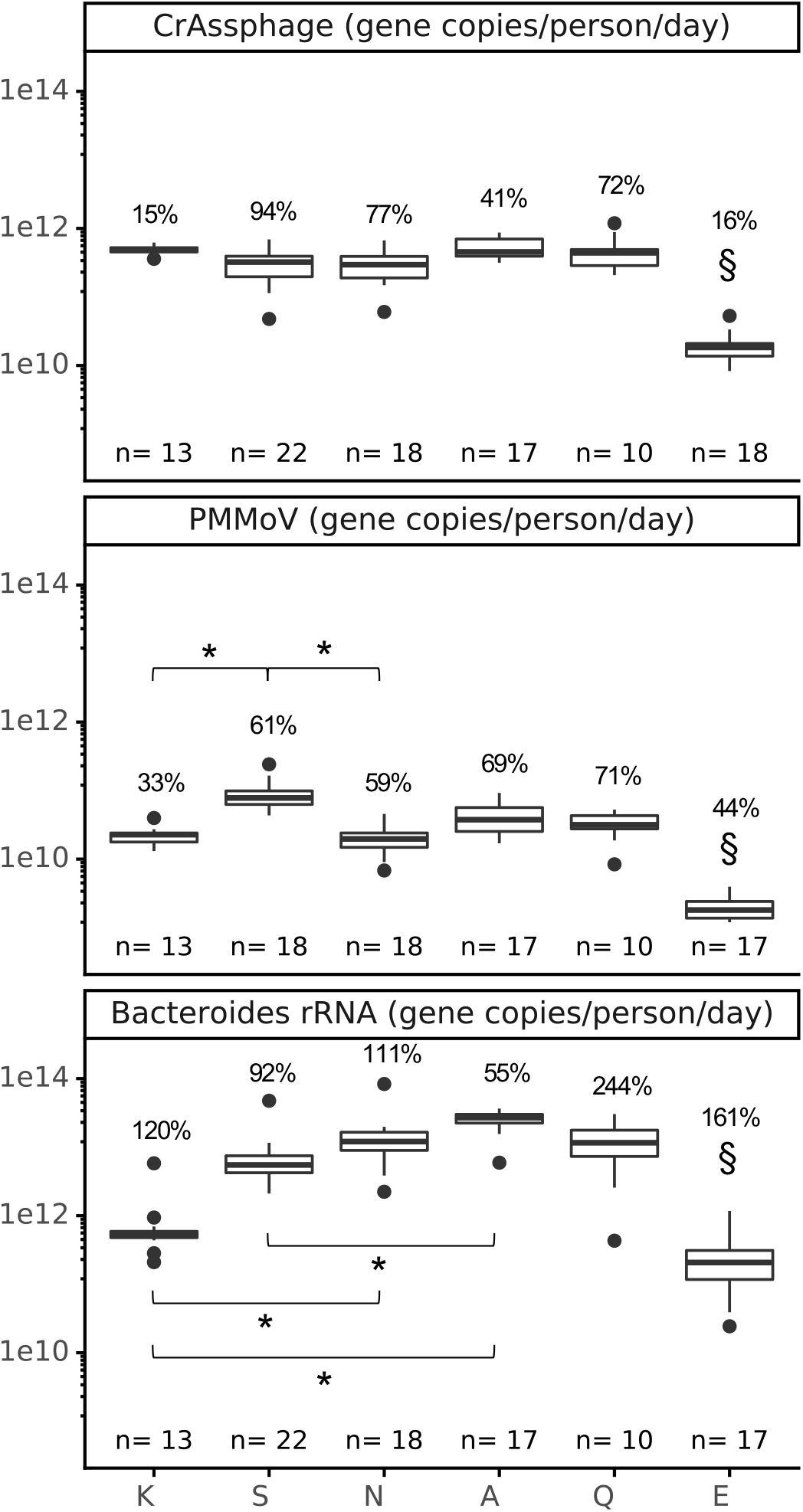
Spatial and temporal variation in crAssphage, PMMoV, and *Bacteroides* wastewater loads. Only one biological replicate per date per location is shown. 18S rRNA results were not included in the figure for consistency of scale due to the wide range in sample values and are included in the SI (**Figure S8**). The temporal variation within each location was assessed as the geometric coefficient of variation, displayed as a percentage above each box. The significance of differences between locations was assessed using a Kruskal-Wallis test with a Bonferroni correction followed by Dunn’s test, where * indicates p < 0.001 for bracketed relationships and § (above location E) indicates p < 0.001 for every pairwise location comparison to E, except p > 0.001 when compared to location K (for crAssphage and *Bacteroides*) and location N (for crAssphage).

### 2.2 Clinical testing and population data

Geospatial vector data of the sewersheds (locations S, K, A, and N) were used to determine the COVID-19 clinical testing data that mapped to each wastewater catchment area (**Table 1**). For all locations, daily new case data correspond to the date that results were reported (result date) for each COVID-19 test. For location K, additional data were available that correspond to the sample collection date and the episode date, defined as the earliest of: (i) the date of first symptoms; (ii) the sample collection date; or (iii) the date the sample was received by the testing lab. Clinical testing data were provided by the corresponding county or open data portal (**Table 1**). Data were masked by public health departments to maintain confidentiality of the contributing population (below 11 new cases per day) and were provided as 7-day (A, S, K) or 14-day (N) moving averages. Masked values were substituted at 5.5 new cases per day for further analysis and plotting. For San Quentin Prison (location Q), unmasked COVID-19 clinical data were obtained from the California Department of Corrections and Rehabilitation open data portal (“CDCR Population COVID-19 Tracking,” n.d.), and instances of zero cases were substituted at 0.5 cases for comparison to masked data in statistical data analysis (**Figure 5**). For clinical data obtained for this study, no IRB was needed because data were either provided masked or were publicly available. More information about masking and population data is provided in **Table 1** and **SI Section E**.

### 2.3 Wastewater sample processing via the 4S method

Wastewater samples were concentrated and extracted following the 4S method (Whitney, 2020; Whitney et al., 2021), with a minor modification: the elution buffer was not pre-warmed; instead, it was added to the column, and the column was heated at 50°C for 10 minutes before centrifugation to collect the eluate. Both RNA and DNA were captured (**Figure S1**). Each extraction batch contained a negative extraction control, and each sample or control was spiked with a surrogate virus control (Bovilis coronavirus; Merck Animal Health, BCoV) and a free RNA control (synthetic oligomer construct, SOC). Because it is not possible to independently quantify the surrogate spike without the influence of extraction efficiency (Kantor et al., 2021), extraction controls were used to assess consistency of extractions rather than recovery. Outlier analysis (alpha=0.05) was conducted for BCoV and SOC Cq values using Grubbs test. No outliers were detected, and all samples tested were considered to have passed this quality control screen. Wastewater sample processing is further described in **SI Section B**.

### 2.4 RT-qPCR plate setup, controls, and data processing

Reverse transcription quantitative polymerase chain reaction (RT-qPCR) was performed on wastewater extract targeting eight sequences: (i) SARS-CoV-2 CDC nucleocapsid gene (N1) assay duplexed with (ii) VetMAX™ Xeno™ Internal Positive Control (Xeno) assay, (iii) crAssphage CPQ_056 (crAssphage) assay, (iv) pepper mild mottle virus coat protein gene (PMMoV) assay, (v) *Bacteroides* 16S ribosomal RNA HF183/BacR287 (*Bacteroides* rRNA) assay, (vi) bovine coronavirus transmembrane protein gene (BCoV) assay, (vii) Synthetic Oligomer Construct T33-21 free-RNA (SOC) assay, and (viii) human 18S ribosomal subunit RNA (18S rRNA) assay (Greenwald, 2021). Reaction conditions (**Table S2**), thermocycling conditions (**Table S3**), and primers, amplicon sequences, and probes (**Table S4**) are included in the SI. Reactions consisted of 20 μL total volume, including 5 μL of RNA extract, TaqMan Fast Virus 1-Step Master Mix (ThermoFisher Scientific), primers, probes, and nuclease-free water. Reactions were completed on a QuantStudio 3 Real-Time qPCR system (ThermoFisher Scientific), where Cq values were determined through automatic thresholding on QuantStudio 3 Design and Analysis Software (v1.5.1). Every plate included samples, no-template controls (NTCs), and standards, each quantified in technical triplicate (qPCR replicates). Individual standard curves (efficiencies ranging from 83.2% to 97.8% and R^2^ ranging from 0.974 to 0.999 for the N1 standard (Twist Bioscience)) were used as a quality control measure (**Table S5**) and later combined into master standard curves (**Table S6**) to calculate quantities (Ahmed et al., 2020c, 2021). A subset of samples were run with no reverse transcription (no-RT) controls for *Bacteroides* rRNA and 18S rRNA, and RNA was found to be multiple orders of magnitude greater than DNA in the samples tested (**Table S7)**. Further details on RT-qPCR materials and no-RT controls are provided in the **SI Section C**.

Raw Cq values that did not amplify or that amplified below the limit of detection were substituted with the Cq value corresponding to half the limit of detection (for N1) or half the lowest point of the master standard curve (for all other assays) (**Table S6**), and then outliers were assessed using a two-sided Grubbs test (alpha=0.05). The N1 qPCR limit of detection (LoD) was calculated by analyzing all RNA standard curves from the study as well as four additional extended triplicate standard curves. The N1 LoD was set at 5 gene copies per reaction, at which point 67% of technical replicates were positive (**Table S8**). Further details on the data processing pipeline are provided in **SI Section D**.

### 2.5 Assessing PCR inhibition via serial dilution and an internal amplification control

To our knowledge, there is no standard methodology for assessing PCR inhibition in raw wastewater samples. We combined two approaches to assess PCR inhibition in raw wastewater samples: a non-competitive internal amplification control (Ahmed et al., 2020b; Nolan et al., 2006; Schrader et al., 2012; Staley et al., 2012) and serial dilution (Graham et al., 2020). The internal amplification control can easily be included in every sample, but cannot detect assay-specific inhibition (Schrader et al., 2012). Serial dilution consumes more resources and risks diluting the target signal below the detection limit, but it more accurately tests the target itself and allows selection of a dilution value that best reduces the impacts of inhibition. Thus, we used the VetMAX™ Xeno™ Internal Positive Control (ThermoFisher Scientific) as a screening tool to select samples for further testing with serial dilution.

For all samples, Xeno RNA was spiked into the reaction mix (**Table S2**), and NTCs were used as an inhibition-free baseline to compare each sample on that plate. Ten samples showed >2 Cq deviation from the baseline and were selected for further inhibition testing (Staley et al., 2012). A dilution series (1x, 2x, 5x, 10x) was performed on these samples, and the duplexed N1 and Xeno assay was repeated. A dilution was chosen by comparing SARS-CoV-2 N1 signal in each dilution to theoretical expectations (based on theoretical doubling per PCR cycle). If diluting the sample led to a 1 Cq difference between actual and expected change in Cq, then the sample at the base dilution was deemed inhibited (Graham et al., 2020). Following the serial dilution test, only three samples required dilution (**Table S9**), and subsequent qPCR results in this study are reported using this chosen dilution. Results from the internal amplification control were inconsistent with inhibition assessed via serial dilution, and we do not recommend the use of Xeno for testing N1 inhibition in future studies.

### 2.6 Data analysis

All data analysis was performed in Python (v3.6.9) using key modules Pandas (v1.1.5), NumPy (v1.19.5), SciPy (v1.4.1), and Plotnine (v0.6.0).

#### 2.6.1 Normalization biomarker analyses

For N1 normalization to biomarkers, N1 (gene copies per liter, gc/L) was divided by the normalization biomarker concentration (gc/L). To calculate flow-scaled biomarker load (gc/person/day), target concentration (gc/L) was multiplied by mean flow for the sampling location (MGD) and a unit conversion factor (liter per million gallons) and then divided by population. Daily flow rate data were not available for S, N, and A (locations upstream of a treatment facility) (**Table S1**), so mean dry weather flow rates (and population) were used to scale data when comparing across locations. We expect that the mean flow rate likely approximates the daily flow rate throughout the study period, but this may not hold true in different locations and seasons.

For comparisons of biomarker concentrations and variation (**Figure 1**), a Kruskall-Wallis test (SciPy v1.4.1) was performed, followed by pairwise Dunn’s tests (scikit-posthocs v0.6.6) to determine statistical differences. Rank correlations between wastewater and case data (**Figure 2**) were calculated as Kendall’s Tau-b coefficients (*τ*; SciPy v1.4.1), a method adapted for left-censored data (i.e., datasets with data below a lower limit of detection) (Wood et al., 2011) because 22% of the data are below the N1 LoD. Correlations were classified as low (*τ* < 0.3), moderate (0.3 < *τ* < 0.5), or high (*τ* > 0.5). Coefficients of variation (CV) were calculated as the arithmetic standard deviation divided by the mean, while geometric coefficient of variation (gCV) was calculated as the geometric standard deviation minus one.

**Figure 2:**
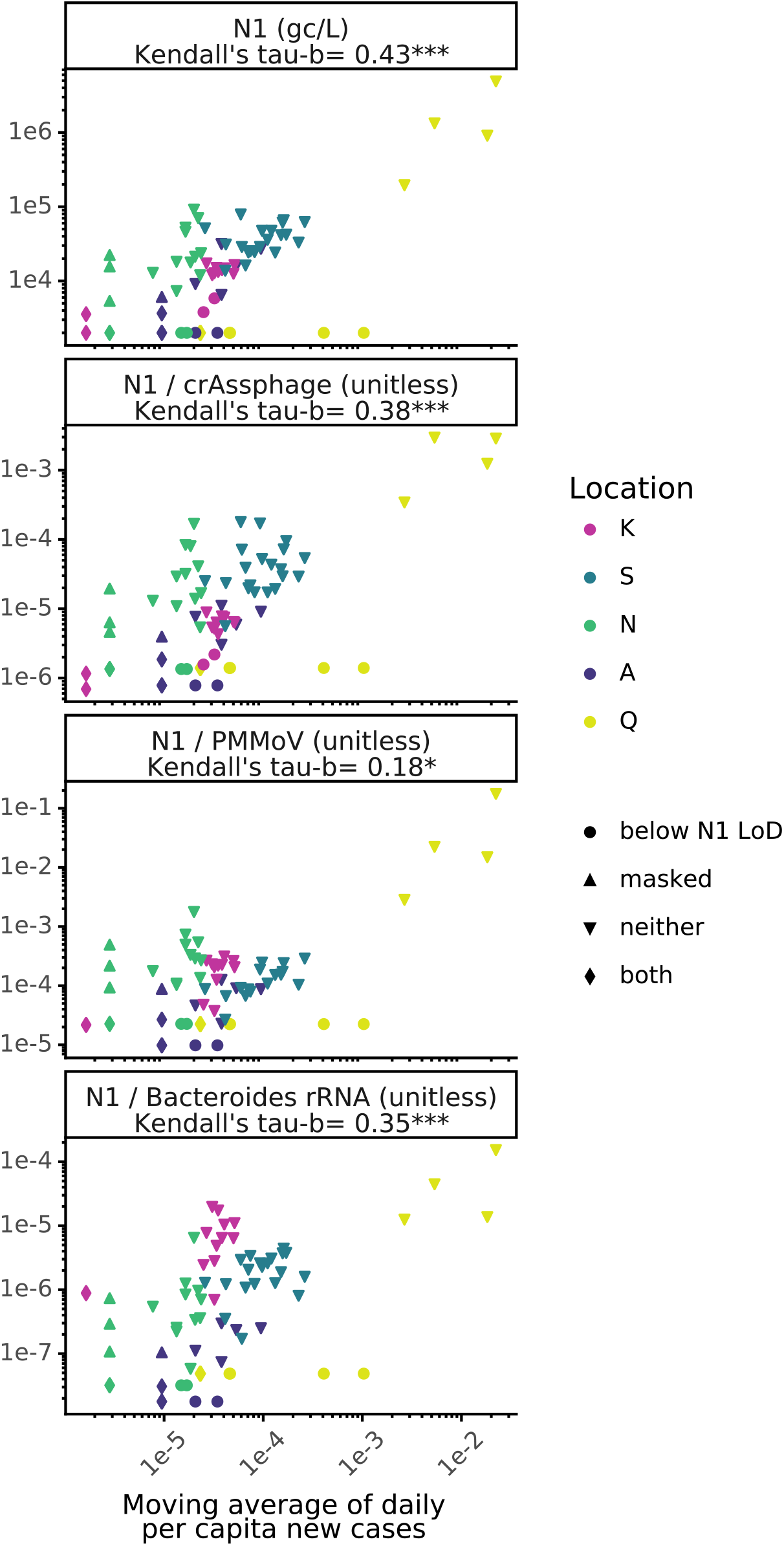
Rank correlations of both unnormalized and normalized wastewater SARS-CoV-2 concentrations with clinical testing data. N1 concentration, and N1 normalized proposed biomarkers are plotted against a seven-day moving average of new cases per capita per day for sample locations K, S, N, A, and Q. Shapes signify whether wastewater samples were below the qPCR limit of detection (LoD) for the N1 assay, associated with masked clinical case values, or both. Significance of rank correlation values in facet titles is indicated by *=<0.05, ***=<0.0001

#### 2.6.2 Assessment of WBE case detection limit

The WBE case detection limit was estimated as follows. The paired wastewater and case data for all sewersheds were combined and sorted from highest to lowest case counts. For each case count, all technical replicates in the wastewater data at and above that point were tallied to determine the cumulative percentage of replicates that amplified in RT-qPCR. Equation 1 was used to fit a logistic function (Kyurkchiev and Markov, 2016) to the dataset (SciPy v1.4.1), where y is the fraction of amplified technical replicates, x is the log_10_(moving average of new cases per person per day), k sets the growth rate of y, and γ sets the inflection point. Zero new cases per capita cannot be represented in a logistic growth model, but in this study, case values of zero were only available for location Q, and these values were substituted as 0.5 cases before the analysis. The COVID-19 per capita case rate that corresponded to 95% cumulative amplification of technical replicates was reported as the estimated WBE case detection limit, and the analysis was repeated with samples where daily per capita cases were provided as masked values.

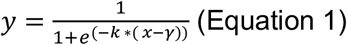

#### 2.6.3 Wastewater trendline smoothing

For wastewater data, any smoothed trendline displayed in a figure was determined using a fitted local regression (Lowess; statsmodels v0.10.2) with bandwidth parameter (α, the fraction of the dataset used for smoothing), set as previously shown (Jacoby, 2000) (**Figures 3 and S2-S5**). Lowess trends of SARS-CoV-2 N1 signal were also visualized as heatmaps to aid in discerning peaks (**Figures S6 and S7**). Full dataset and associated code are available through GitHub (https://zenodo.org/record/4730990#.YIxkrqlKgUo).

**Figure 3:**
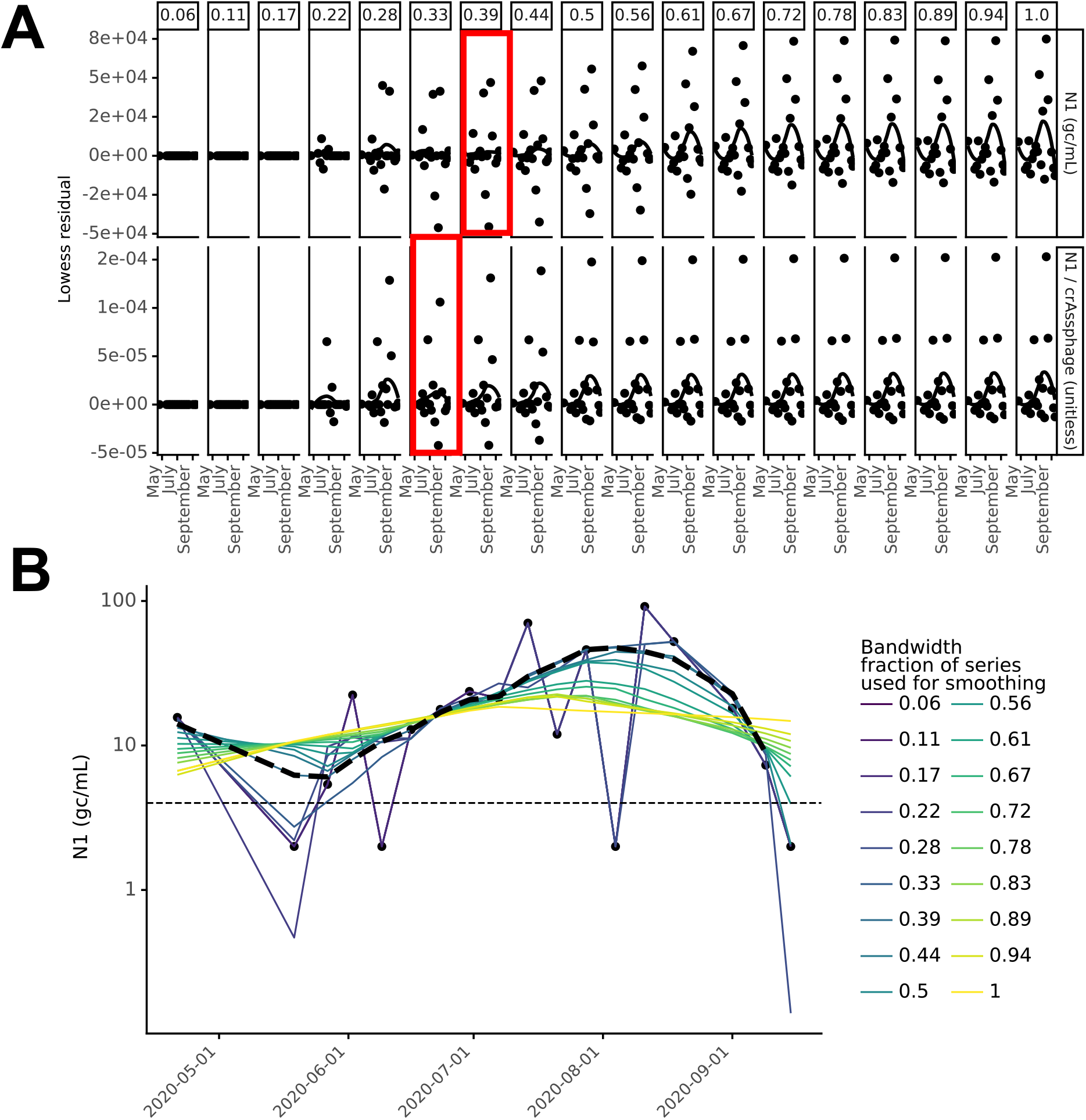
Example of Lowess bandwidth parameter selection process (Location N) (A) Residual plots for Lowess bandwidth parameter (α; column labels) determination for location N where the bandwidth parameter increases from inclusion of 1 data point (far left) to inclusion of all data points (far right) in each local regression for unnormalized N1 (top) and crAssphage-normalized N1 (bottom). The value of α that minimized the residual was selected (red boxes). (B) Visualization of how bandwidth parameter affected the Lowess trendline for location N. Black dashed line indicates the resulting Lowess trendline when α=0.39.

## 3. RESULTS

Raw wastewater was collected weekly from April to September 2020 at six locations (**Table 1**). The resulting dataset includes 91 samples (155 including biological replicates), analyzed for SARS-CoV-2 and four potential normalization biomarkers (crAssphage, PMMoV, *Bacteroides* rRNA, and human 18S rRNA) and paired with geocoded clinical testing data. This dataset was generated from the San Francisco Bay Area in separate sanitary sewer systems during a period with minimal rainfall (see Methods), which naturally controlled for variability in wastewater strength due to precipitation. Thus, we expected the concentrations of the measured normalization biomarkers to be relatively stable. Additionally, geocoded clinical testing data included a range of per capita COVID-19 case rates that varied by location.

### 3.1 CrAssphage and PMMoV were the most consistent biomarkers

A subset of samples from all locations were used in experiments (**Figure 1**) comparing crAssphage (98 unique samples, 153 biological replicates), PMMoV (93 unique samples, 95 biological replicates), *Bacteroides* rRNA (97 unique samples, 99 biological replicates) and 18S rRNA (40 unique samples, 41 biological replicates) as biomarkers for normalization to fecal content. All normalization biomarkers were detected at high concentrations (**Table S10**) in all samples tested, except for 18S rRNA, which was inconsistently quantifiable (**Figure S8**). Flow rates and chemical wastewater parameters (TSS, BOD, COD, cBOD) were not consistently measured by utilities (**Table S1**); thus, robust comparisons of physicochemical biomarkers could not be made. In the absence of daily flow rate data, we used mean flow rate to scale wastewater biomarker concentrations by per capita wastewater flow for each sewershed to account for differences across sewersheds (**Figure 1**). Mean per capita flow rates were similar for all locations except Q (a facility; **Table 1**), generally resulting in little change after flow-scaling. For this reason, flow-scaling was applied to compare biomarkers, but unnormalized SARS-CoV-2 N1 concentrations are used as a baseline in later analyses.

An ideal normalization biomarker would have minimal spatial variation in per capita shedding rates and minimal temporal differences in wastewater loads when flow rates are stable, as they were in this study. Two methods were used to evaluate biomarker variability (**Figure 1**): (i) comparing per capita biomarker loads (gene copies/person/day) to assess differences in observed shedding by location; and (ii) evaluating the temporal variation of loads for each location. Consistent with recent studies (Ahmed et al., 2020d; D’Aoust et al., 2021b), crAssphage and PMMoV were the least variable biomarkers across locations and over time (mean gCV_crAssphage_=59% and mean gCV_PMMoV_=56%, not statistically different (p>0.05)). In contrast, *Bacteroides* rRNA displayed more variability both spatially (**Figure 1**) and temporally (mean gCV=130%), and 18S rRNA varied dramatically (mean gCV=500%) (**Figure S8**).

CrAssphage, PMMoV, and *Bacteroides* rRNA were quantifiable in all samples tested, but 18S rRNA was below the LoD for 24% of samples. Furthermore, 18S rRNA was the only biomarker that amplified in the extraction negative controls (75%) at similar levels as the samples (p>0.05; **Figure S9**), which suggests that this target could be a common laboratory contaminant. Based on our findings that 18S rRNA was frequently detected in negative controls, inconsistently detected in sewage, and had high spatial and temporal variation in per capita shedding, we conclude that human 18S rRNA is not suitable as a normalization biomarker to adjust for fecal content.

We suspected that lower biomarker concentrations (**Figure 1**), frequently undetected 18S rRNA (**Figure S9**), and high variability in N1 signal in location E samples reflected RNA degradation because: (i) location E is the largest sewershed in the study (i.e., longer residence time allowing for signal degradation); and (ii) some samples from this site thawed during transportation back to the lab (resulting in an additional freeze-thaw cycle, which could degrade RNA (Ahmed et al., 2020c; Coryell et al., 2020)). Data from sewershed E was not used for subsequent analyses because of the uncertainty surrounding the integrity of these samples.

### 3.2 SARS-CoV-2 N1 and clinical testing data were correlated and some normalization biomarkers maintained this relationship

The SARS-CoV-2 N1 concentration in wastewater was moderately correlated to daily per capita clinical cases when aggregated across all locations (*τ*=0.43, p<0.0001; **Figure 2**). However, there are several limitations to assessing correlation between clinical testing data and wastewater data. First, clinical testing data do not necessarily represent true incidence because of biases associated with testing. Even if clinical and wastewater testing data correspond to the same date, fecal shedding could peak before symptom onset, which would impact the correlation unless the correct time offset is applied to reflect this discrepancy (Hoffmann and Alsing, 2021). Additionally, for this analysis, clinical and wastewater testing data would ideally both correspond to the sample collection date (as opposed to the result date) to remove lag introduced by test result turnaround time (see section 3.3 for more information). However, often only one date was available for clinical testing. For example, in this analysis, the only date associated with the daily per capita cases from most locations (all but K) was the date that the testing results were provided (result date), while the date associated with the SARS-CoV-2 N1 wastewater signal was the sample collection date.

Despite these potential limitations, correlation to daily per capita COVID-19 cases was used as a metric to assess the effect of normalization to biomarkers (crAssphage, PMMoV, and *Bacteroides* rRNA). Moderate, significant correlations were observed with COVID-19 daily per capita cases when SARS-CoV-2 N1 was unnormalized and normalized by crAssphage or *Bacteroides* (*τ*_unnormalized_=0.43, *τ*_crAssphage_=0.38 and *τ*_*Bacteroides*_=0.35, p<0.0001; **Figure 2**); however, normalization did not strengthen the correlation compared to unnormalized signal. Conversely, PMMoV normalization produced only a weak correlation of 0.18 (p < 0.05) (**Figure 2**). Analysis was performed with and without samples that were below the limit of detection and produced similar results (**Table S11**). Of the normalization biomarkers tested, crAssphage had the lowest variability and also maintained significant and moderate correlation with clinical testing data, so we included it in subsequent analyses alongside unnormalized concentrations.

The correlation analysis was repeated with data separated by location to determine whether locational dependencies affect the relationship between wastewater and clinical testing data as well as the performance of normalization strategies. Locations with at least 75% of data above the N1 qPCR LoD (Locations K, N, and S; **Table S12**) were included in this analysis. Only location K had significant correlations with clinical testing data, both with and without crAssphage normalization (*τ*_unnormalized_=0.5 and *τ*_crAssphage_=0.43, p<0.05; **Figure S10)**. This finding suggests locational dependencies (e.g., differences in extent of clinical testing, sewer system residence times, etc.) affect the correlation with clinical testing data. Additionally, the location-specific analysis was repeated including only samples with detectable SARS-CoV-2 N1 signal, and the results were no longer statistically significant (**Figure S10)**. This finding is likely influenced by both the limited sample size and values below the N1 qPCR LoD that affect rank correlations.

In addition to the limitations in clinical testing data mentioned at the beginning of this section, there are several explanations for why wastewater signal at locations S, N and K did not significantly correlate with clinical testing data after removing values below the LoD: (i) the daily per capita cases in the population were at or below the WBE case detection limit of the wastewater data; (ii) the daily per capita cases that were masked by public health departments for patient privacy imparied the rank correlation analysis by left-censoring the clinical testing data; (iii) the wastewater signal did not vary enough over the time of sampling to establish rank. The possibility that the wastewater signal leads the clinical testing data was subsequently tested for locations K, N, and S (i.e., correlations were examined for zero-, one-, and two-week offsets); however, location K was the only location with significant correlation between wastewater and clinical testing data for any lead time tested (**Figure S10)**.

### 3.3 Impact of the date associated with clinical testing data on lead time in wastewater surveillance at location K

The time for laboratories to process samples and return results (testing turnaround time) affects the potential for wastewater surveillance to provide lead time over clinical surveillance. In general, clinical testing data correspond to either the date the sample was collected or the date the results were returned. The ideal date to use for informing public health decisions would be the result date, to include differences between clinical and wastewater testing turnaround time in the analysis. Alternatively, sample collection dates should be compared to understand the timing of the underlying biological mechanisms that result in a positive wastewater signal (onset and duration of fecal shedding) and positive clinical test (onset and duration of nasopharyngeal shedding). Onset and duration of symptoms may influence the timing of the clinical test (sample collection date), depending on whether testing is routine or only available to symptomatic individuals. Hence, the ideal date to use for comparison of wastewater and clinical testing data differs depending on the goals of the comparison. The clinical testing data for location K included sample collection date, result date, and episode date (the earliest date associated with the case), allowing us to assess the correlation between case data and wastewater data with and without clinical testing turnaround time. Episode date was frequently the same as the sample collection date, unless a patient reported symptoms prior to test date (**Figure S11**). It should be noted that wastewater testing data correspond to the sample collection date because all samples were processed retroactively in this study. Routine wastewater testing turnaround time can be 1-3 days but could vary depending on sample transport and laboratory methods (“Covid-WEB,” n.d.).

To test the influence of the date associated with clinical testing, we repeated correlation analysis for location K (**Figure S11**). The wastewater testing data (sample collection date) correlated with the clinical testing data by episode date (*τ*_episode,unnormalized_=0.56, *τ*_episode,crAssphage_=0.54, p<0.01) and sample collection date (*τ*_collection,unnormalized_=0.59, *τ*_collection,crAssphage_=0.62, p<0.01) without a lead or lag. When the result date was used for clinical testing data, the strongest correlation with wastewater data was associated with a two-week lead time (unnormalized N1 concentration) or one-week lead time (N1 normalized to crAssphage; **Figures S10** and **S11**). When values below the N1 qPCR LoD were removed, wastewater data were no longer significantly correlated with episode date-associated clinical data, but the strongest correlations for the other date associations remained significant. This analysis is limited because of the small dataset, but the methodology presented here can be used to assess the lead time provided by wastewater surveillance with larger data sets and with wastewater data processed contemporaneously with decision-making.

### 3.4 The Lowess bandwidth parameter affected wastewater data trend interpretation

Variation in wastewater SARS-CoV-2 N1 signal from sources other than variation in true incidence or prevalence (e.g., noise introduced during sample collection, processing, etc.) can obscure temporal trends. Smoothing techniques can be used to visually distinguish temporal trends from noise. Similar to the choice of the number of days included for each average calculation for moving averages (window), Lowess requires selection of the fraction of the whole time series that is used for each local regression calculation (bandwidth). We employed a method to set the bandwidth parameter systematically based on residuals (Jacoby, 2000) independently for each location. The bandwidth was increased stepwise, beginning with inclusion of one point in each local regression calculation and ending with inclusion of all points (α=1). For each bandwidth value, the residuals were calculated and plotted by date, and a Lowess trendline with α=1 was fit to these residual plots to monitor residual trends as the bandwidth varied. Finally, the maximum bandwidth value was selected for which the residuals visually maintained horizontal Lowess trendlines (see **Figures 3 and S2-S5**).

As an example, for unnormalized and crAssphage-normalized SARS-CoV-2 N1, bandwidth parameters of 0.39 and 0.33 were respectively chosen for location N (**Figure 3 A**). This process was repeated for all locations, and bandwidths in the range of 0.25-0.6 were selected based on the optimization procedure (see **Figures 3 and S2-S5**). To assess the impact of bandwidth on SARS-CoV-2 N1 signal interpretation, Lowess was performed for all locations sampled and for all possible bandwidths (see **Figures 3 B and S2-S5**). The bandwidth parameter influenced the overall temporal trends of wastewater data for some locations (N and A; **Figures 3 and S5**). For example, at location A (**Figure S5**), a bandwidth of 1 resulted in a gradual increase in SARS-CoV-2 N1 signal during sampling, while a bandwidth of 0.73 resulted in a peak around July. However, for location K (**Figure S2**), all bandwidths resulted in trends that would have similar interpretations. These results illustrate that choice of bandwidth could have implications for interpreting WBE data and informing COVID-19 response strategies, and systematic methods should be used to select the appropriate bandwidth.

### 3.5 Wastewater and clinical data had similar overall trends regardless of normalization, with notable exceptions

To assess the impact of crAssphage normalization on SARS-CoV-2 N1 temporal trends, we compared unnormalized and crAssphage-normalized Lowess trendlines (**Figure 4**). We found that crAssphage-normalized trends were similar to unnormalized trends for three of the locations (K, N, and A) but had differences in overall trend for locations Q (**Figure 5**) and S (**Figure S12**).

**Figure 4:**
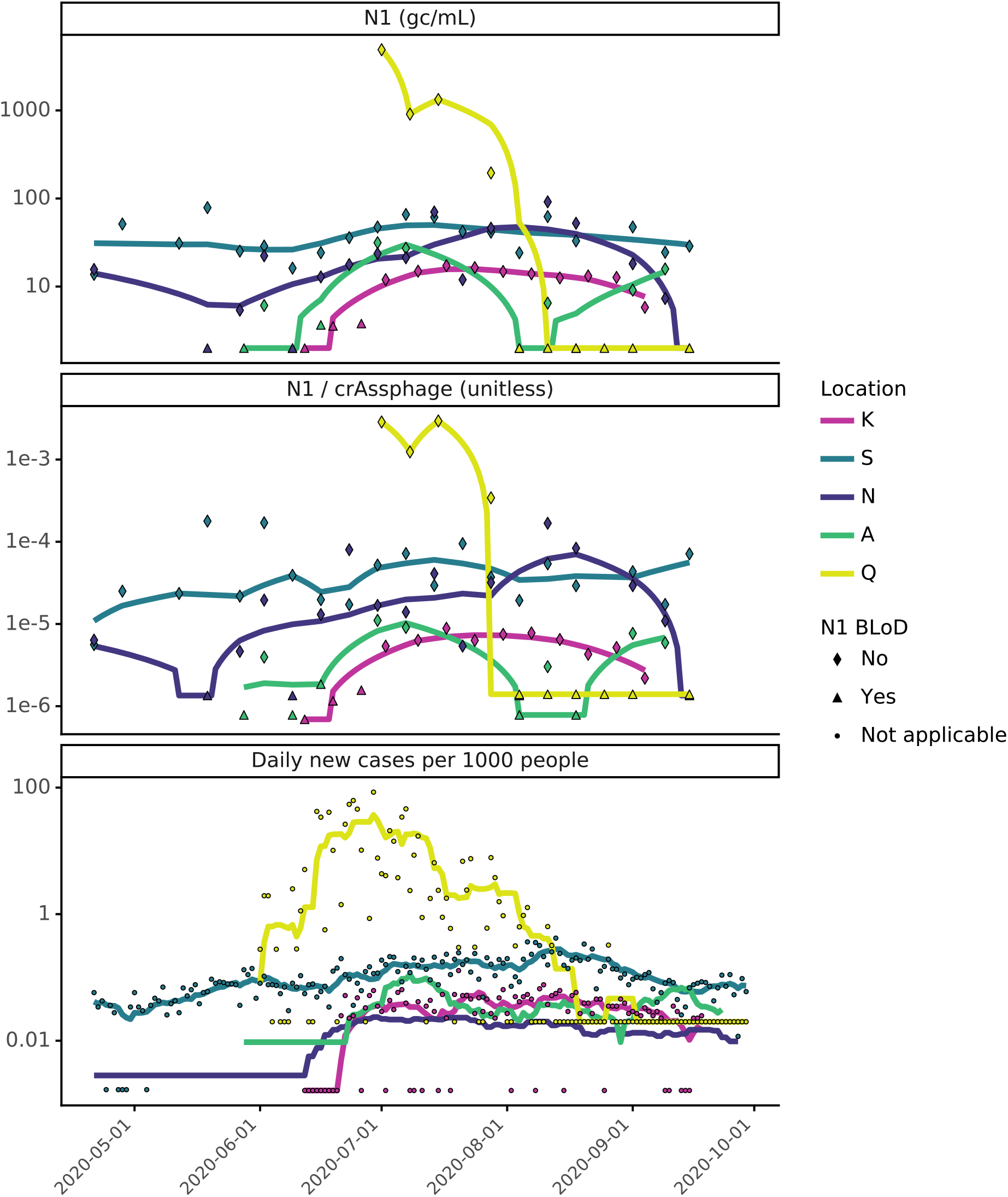
Comparison of wastewater SARS-CoV-2 N1 to geocoded COVID-19 clinical testing results from May to September 2020. Wastewater SARS-CoV-2 N1 signal is compared as unnormalized (top) and crAssphage-normalized (middle), where lines are the most optimal Lowess trendlines. COVID-19 clinical testing results are the daily per capita COVID-19 cases, where lines are the fourteen-day moving average (location N) or seven-day moving averages (all other locations) (bottom). Heatmap visualization of the unnormalized N1 trendlines is included in the SI (**Figures S6 and S7**) and visualization of sewersheds by location can be found in **Figure S12**.

**Figure 5:**
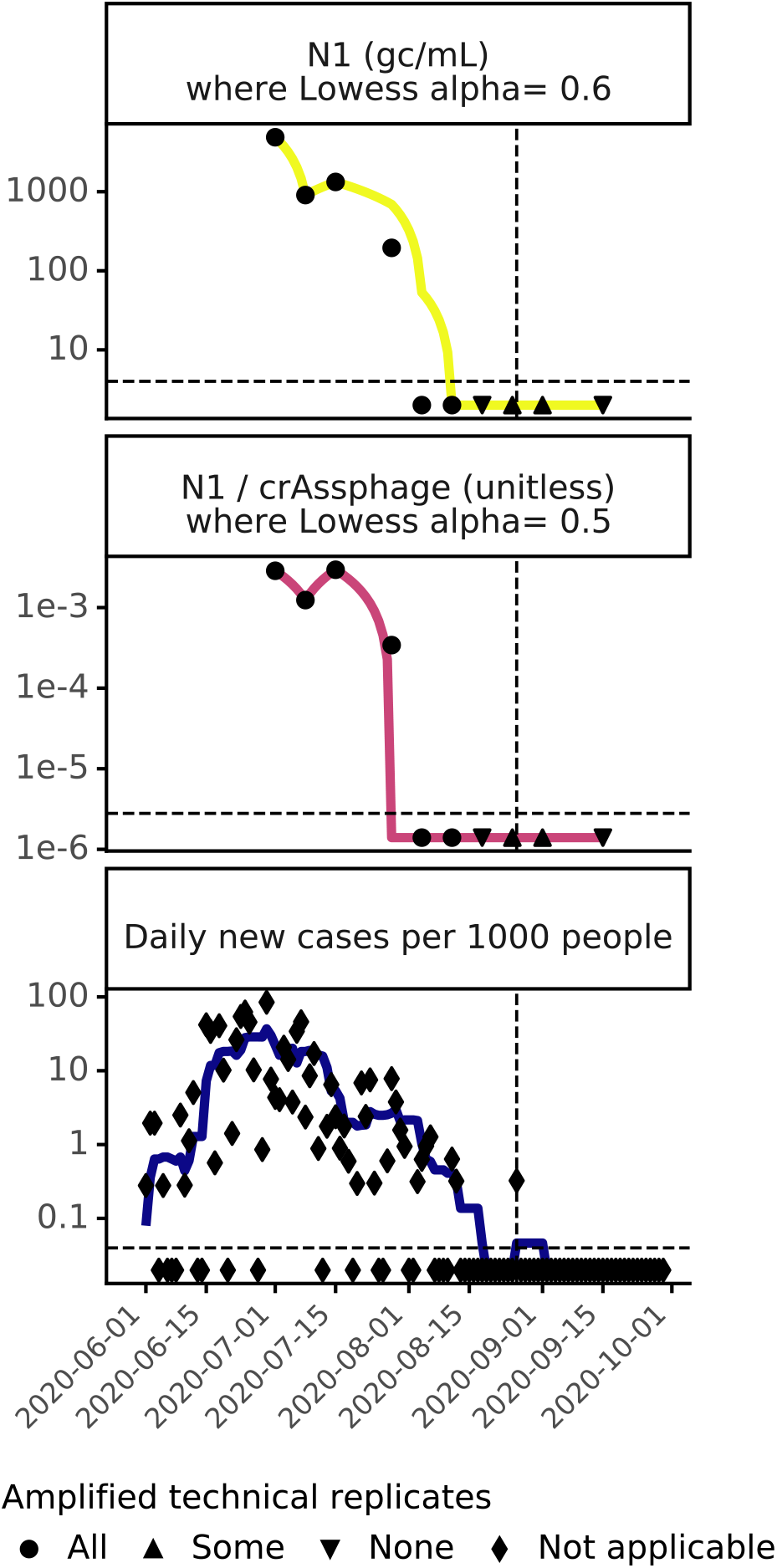
Comparison of wastewater and clinical data at location Q from June to September 2020, where symbols indicate how many technical replicates amplified during qPCR. **Wastewater data : (top)** unnormalized and **(middle)** crAssphage-normalized SARS-CoV-2 N1 signal in wastewater, where the horizontal dashed line indicates the limit of detection, and trendlines are the most optimal Lowess trendline (**Figure S4**). **Clinical data (bottom):** daily per capita COVID-19 cases, where the horizontal dashed line indicates 1 case in 1000 people. Vertical dashed lines indicate August 26th, the only date after August 12th when a new COVID-19 case was detected at location Q through clinical surveillance.

Discrepancies are concerning because they could have implications for pandemic response. We note that the trend in location K, for which biological replicates were processed routinely, was the least impacted by bandwidth or normalization (**Figures S2 and S12**). Larger datasets with more frequent sampling and processing of biological replicates, would give single points less influence over the trend.

Relative spatio-temporal trends in clinical and wastewater testing results were compared across sampling sites (**Figures 4, S6, and S7**). In general, clinical and wastewater data at all locations paralleled one another, with San Quentin prison (Q) showing the highest COVID-19 burden across locations. Due to a COVID-19 outbreak, location Q had a maximum that was 53 times (SARS-CoV-2 N1 4.89 x 10^3^ gene copies/mL), 17 times (crAssphage-normalized SARS-CoV-2 N1 ratio 2.9x 10^−3^), and 203 times (∼85 new cases per 1000 people on 6/29) higher than the highest value at the sewershed scale. There were a few discrepancies between clinical and wastewater trends (heatmap visualizations in Figures S6 and S7 highlight discrepancies in peaks). For example, at location N, there may have been clinical undertesting, based on the peak in wastewater data in August (**Figures 4 and S6**) and higher SARS-CoV-2 signal in wastewater at location N (relative to other locations) than represented by the clinical data (**Figures 4 and S7**).

### 3.6 The WBE case detection limit was estimated to be 2.4 COVID-19 cases per 100,000 people

Quantifying the minimum per capita new COVID-19 cases in a sewershed at which there is reliable detection of SARS-CoV-2 N1 in wastewater (WBE case detection limit) is important for gauging the utility of COVID-19 WBE when the true incidence is low. This WBE case detection limit depends on the detection limit of the wastewater measurement (i.e., the methods used to store, concentrate, extract, and measure SARS-CoV-2 RNA in wastewater) and the accuracy of the clinical testing data available. To estimate the WBE case detection limit in a way that is replicable across studies, the cumulative percentage of amplified technical replicates of the wastewater data for inversely-ranked daily per capita COVID-19 cases was fit to a logistic growth model (without samples associated with masked case values; see Methods). When COVID-19 case rates equaled or exceeded 2.4 daily cases per 100,000 people, 95% of wastewater technical replicates amplified via RT-qPCR for N1 (**Figure 6**). Other researchers have used non-cumulative methods to estimate the WBE case detection limit by calculating the percent of amplified wastewater replicates for each case value (Wu et al., 2021). This method requires repeated wastewater measurements associated with each possible clinical case value or range of case values (i.e., bins). Otherwise, the percent of amplified technical replicates is limited, as was the case in this study where only one biological replicate was often associated with each case number (**Figure S13 A**). Ideally, all data would be unmasked when applying this method. To verify that the masked clinical data did not affect the estimated WBE case detection limit, the process was repeated with masked values, and the estimate was similar (2.2 cases in 100,000 people; **Figure S13 B**). These limits are within the theoretical range possible (Hart and Halden, 2020b) and similar in magnitude to previous findings of 10 in 100,000 (Hata and Honda, 2020) and 13 in 100,000 (Wu et al., 2021).

**Figure 6:**
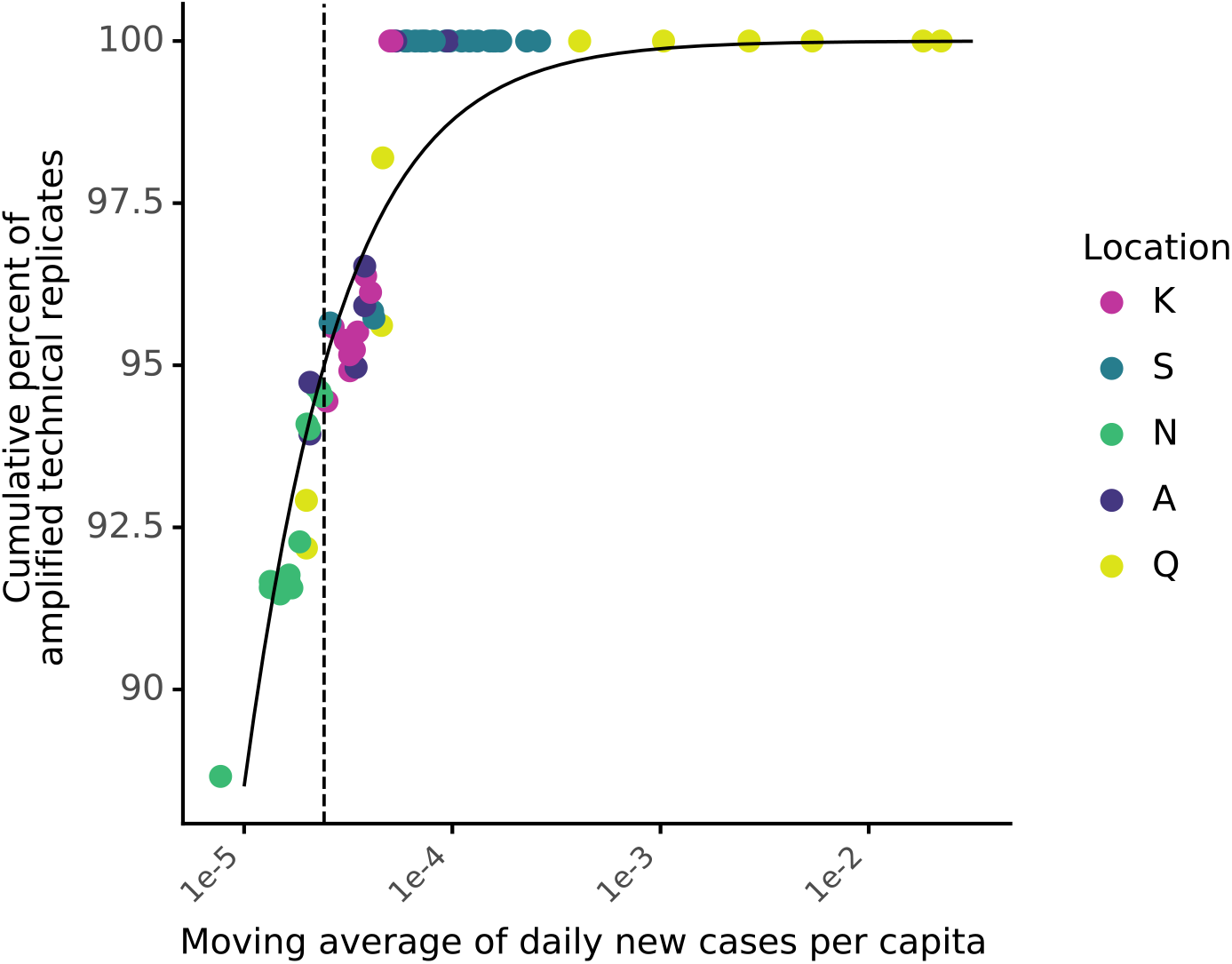
Estimated minimum number of COVID-19 clinical cases needed for reliable detection of SARS-CoV-2 RNA in wastewater. The cumulative percentage of amplified wastewater technical replicates was calculated by ranking the moving averages of daily per capita cases (x-axis) from highest to lowest and calculating the fraction of qPCR replicates that amplified cumulatively (y-axis) for each value of x. The dashed line represents the daily new cases per capita value above which 95% of wastewater technical replicates amplified (2.4 cases in 100,000 people).

Based on the contributing population of each sewershed in this study, the WBE case detection limit translates to 11.6 cases for K, 11.3 cases for S, 3.3 cases for N, 2.0 cases for A, and 0.1 cases for Q. Therefore, at location Q, one case should be reliably detected based on results from this analysis, which could be tested observationally because only one new case was detected by clinical testing after August 12^th^ (**Figure 5)** with the test results provided on 8/26. When wastewater sampling was completed on 8/25 and 9/1, a weak positive signal was observed (one technical replicate amplified). These results support that wastewater testing can detect clinical cases in facilities with a single case; however, to capture a strong positive signal from one case at a facility, higher frequency sampling is recommended. A possible explanation for why a single case may still go undetected, despite being above the WBE case detection limit, is that wastewater surveillance relies on the autosampler aliquots capturing the feces from each infected individual, which becomes less likely when there are fewer infected individuals and wastewater has less mixing prior to the sampling location. These results are promising for the application of WBE in facilities, where it is highly resource-intensive to conduct routine clinical surveillance testing frequently enough to detect a single case in time to enact preventative measures. More work is needed applying WBE for SARS-CoV-2 across a broader set of facilities.

## 4. DISCUSSION

### 4.1 Validation and potential use scenarios of SARS-CoV-2 wastewater testing

During the COVID-19 pandemic, both the methodological research for SARS-CoV-2 testing in wastewater and the application of WBE have been occurring simultaneously. For COVID-19 WBE to be useful for public health decision-making, public health officials need to be confident that the resulting SARS-CoV-2 signal reflects COVID-19 trends in the contributing population. Despite limitations in clinical testing data and the potential lag in wastewater trends, assessing correlations between clinical and wastewater testing data can help validate WBE (Xagoraraki and O’Brien, 2020). Moderate correlations with clinical data observed in this study (*τ*=0.43) support that trends in wastewater surveillance data reflect trends in COVID-19 disease occurrence. Wastewater data paired with clinical data can be a more robust public health surveillance strategy compared to either method alone, both for sewershed-scale and facility-scale surveillance applications. In some settings, wastewater testing may be a less resource-intensive way to implement population-scale surveillance, and policymakers will need to balance allocation of resources to each approach.

A critical question for public health decision-making is how much early warning WBE can provide ahead of clinical testing, which could allow more timely public health responses to slow COVID-19 outbreaks. However, lead time is difficult to measure. Biologically, the time between onset of fecal shedding and nasal shedding is unclear (Benefield et al., 2020; Walsh et al., 2020). Practically, lead time depends on testing turnaround time and frequency of sampling for both wastewater and clinical testing. For example, clinical testing capabilities can increase the lead time of wastewater data if patients are only tested after symptom onset and can decrease the lead time if asymptomatic and symptomatic individuals are regularly screened with rapid turnaround time. Ideal assessments of wastewater data lead time due to biological mechanisms would not include turnaround time, whereas assessments of the performance of clinical and wastewater laboratories for public health action and practical limitations would include turnaround time. Although other studies observed lead time for wastewater data over clinical data starting on the order of days (D’Aoust et al., 2021a; Nemudryi et al., 2020; Peccia et al., 2020, the weekly sampling in our study could explain why no lead time was determined when the sample collection date was used for both wastewater and clinical testing data (**Figure S14**). However, the impact of clinical testing strategy (i.e., only screening symptomatic individuals) could also be affecting this result. We could not directly compare wastewater and clinical result dates in this retroactive study, but when clinical data were associated with the result date and wastewater data were associated with sample collection date, lead time of 1-2 weeks was observed (**Figure S14**). Other researchers have observed lead time in wastewater data of up to three weeks (Ahmed et al., 2021; Medema et al., 2020), and our results reflect a similar range in possible lead times (0-2 weeks) depending on which date is associated with the clinical data.

At the sewershed scale, the benefit of WBE to public health extends beyond early warning. Discrepancies between wastewater testing data and clinical testing data trends from early in the time series at location N (April-July 2020; **Figure S12**) could be used to infer clinical undertesting, which is supported by lower testing capacity in this time frame (**Figure S15**). Although pairing COVID-19 clinical testing data with wastewater SARS-CoV-2 data can generate new insights for public health decision-making, it can be challenging in practice. Pairing wastewater SARS-CoV-2 data with geocoded COVID-19 clinical testing data required collaboration between academics, wastewater treatment facility representatives, and public health officials. These collaborations may be particularly difficult at sewershed-scale, where multiple public health department jurisdictions overlap (e.g., location N). Partnerships for data sharing between agencies are critical to support ongoing wastewater-based epidemiology for SARS-CoV-2 and other pathogens.

At the facility scale, monitoring raw wastewater for SARS-CoV-2 might be particularly useful for early detection of COVID-19 outbreaks. San Quentin Prison (location Q) had a COVID-19 outbreak during the study period after a transfer from the California Institution for Men (Cassidy and Fagone, 2020), where, at its peak, 47% of the population had active cases. The maximum SARS-CoV-2 N1 concentration (4.89 x 10^3^ gene copies/mL) was higher than any sewershed sampled in this study and among the highest values we found in the literature for N1 in raw wastewater (Gerrity et al., 2021; Gonzalez et al., 2020; Medema et al., 2020; Randazzo et al., 2020b; Wu et al., 2020; Wurtzer et al., 2020), despite regular clinical testing (**Figure S15**). Prison conditions cause incarcerated people to be particularly susceptible to respiratory disease outbreaks, and maintaining safety in prisons requires deliberate planning and coordination by correctional institutions (e.g., coordination with local public health systems to develop pandemic response plans, coordination of transfers between institutions, etc.) (Montoya-Barthelemy et al., 2020). Furthermore, the health of incarcerated people is linked to the health of the community, and incorporating correctional institutions into community safety plans will help ensure better protection against COVID-19 for everyone (Montoya-Barthelemy et al., 2020). Once protective measures are implemented, WBE may be useful to monitor prisons and other high-risk facilities (e.g., skilled nursing facilities, homeless shelters, etc.), especially where clinical testing is not available or routine.

### 4.2 Approaches for translatable WBE

#### 4.2.1 Normalization of wastewater targets to adjust for fecal content

Results from this study suggest that PMMoV, *Bacteroides* rRNA, and 18S rRNA were less promising normalization biomarkers than crAssphage. While PMMoV was present in high and stable concentrations, the diet-dependency (Symonds et al., 2019) and large range in concentrations in the literature (six orders of magnitude; **Table S10**) remain concerns for its use over longer time scales and across larger geographic regions. Normalization to PMMoV resulted in the weakest significant correlation to clinical testing data of the biomarkers tested (*τ*=0.18, p<0.05), in contrast to other studies that found normalization to PMMoV improved correlation with clinical data (D’Aoust et al., 2021b; Wu et al., 2020). *Bacteroides* rRNA loads varied more spatially and temporally than crAssphage or PMMoV in this study (**Figure 1**), but *Bacteroides*-normalized SARS-CoV-2 N1 had a moderate correlation with clinical testing data (*τ*=0.35). While measurement of *Bacteroides* rRNA gene in wastewater has been commonly applied for fecal source tracking and *Bacteroides* rRNA has been targeted to increase assay sensitivity (D’Aoust et al., 2021b; Feng et al., 2021), to our knowledge, no prior raw wastewater values have been reported in the literature for *Bacteroides* rRNA (**Table S10**). Similarly, no values were found in the literature for 18S rRNA concentrations in raw wastewater (**Table S10**). In this study, 18S rRNA signal displayed a wide range in concentrations and consistently amplified in negative extraction controls. Furthermore, 18S rRNA was less stable in wastewater than SARS-CoV-2 RNA and nonenveloped viruses (e.g., crAssphage and PMMoV), which is consistent with previous studies (Whitney et al., 2021; Wurtzer et al., 2020). Therefore, we do not recommend 18S rRNA use as a normalization biomarker. In comparison to all of the biomarkers tested, crAssphage had low spatial variability (i.e., the fewest locations with statistically different loads; **Figure 1**) and temporal variability (gCV=59%; **Figure 1**). Additionally, normalized SARS-CoV-2 N1 correlated with daily per capita COVID-19 cases (*τ*=0.38). Although crAssphage concentrations in the literature had a wide range (six orders of magnitude; **Table S10**), they were consistent across locations in this study. Based on this dataset, crAssphage remains a promising endogenous normalization biomarker for broader WBE applications.

Although a standardized approach would facilitate comparisons across studies, the ideal normalization strategy may be situationally dependent. For example, in this study, for some locations, crAssphage-normalization did not have a major impact on general spatio-temporal trends and did not improve correlations to clinical data compared to unnormalized signal, likely due to the lack of precipitation or changes in flow throughout the study period. Several factors should be considered when deciding whether to normalize to a biomarker or report unnormalized concentrations. First, adding another assay introduces additional analytical variation that could outweigh the benefits of biomarker normalization in some settings (Feng et al., 2021). An additional consideration is ensuring methods compatibility with the WBE target and normalization biomarker. Ideally, normalization to an endogenous biomarker would account for losses in target signal during residence time in sewers, sample storage, and laboratory processing, but the ideal biomarker for fecal content may not be the best surrogate for the target of interest. For example, crAssphage is not expected to be a good surrogate for SARS-CoV-2 stability, partitioning, and extraction (Ye et al., 2016), and as a DNA virus, crAssphage may be incompatible with some extraction methods used for SARS-CoV-2 RNA. Other controls (e.g., endogenous biomarkers, recovery controls) and modeling may be applied to improve measurement accuracy and translate results across labs and methods, although there are challenges associated with these corrections (Kantor et al., 2021). Degradation modeling with target-specific decay constants (Ahmed et al., 2020a) and sewershed-specific parameters could assist in correcting for degradation or determining sample integrity, but no comprehensive approach for this correction exists.

#### 4.2.3 A systematic approach for data smoothing (Lowess)

In general, public health decisions are based on temporal trends in disease burden, not individual data points, but trends in wastewater and clinical data can be difficult to visually distinguish, especially when available resources constrain sampling frequencies. Applying Lowess to wastewater data, we found that the value of one parameter could influence the trend visualization such that the same dataset could lead to different public health responses (**Figures 3 and S5**). Based on our analysis, the bandwidth parameter for Lowess should be determined for each sewershed sampled. Lowess with a systematically chosen bandwidth could be used to smooth trendlines and minimize the loss of temporal resolution. The method presented here could be applied in retrospective analysis or in real-time analysis completed as part of wastewater public health surveillance programs. For real-time applications, the bandwidth parameter could be selected using a subset of data, and the residuals plot could be frequently checked to ensure no new residual patterns emerge over time that could obscure the smoothed trend.

#### 4.2.4 A systematic approach to estimate a WBE case detection limit

In addition to data smoothing, we developed an approach for identifying a WBE case detection limit that can be applied systematically to studies using PCR-based methods. We applied this analysis to SARS-CoV-2 N1 signal in wastewater and found that the daily new clinical cases at which wastewater surveillance could reliably detect clinically diagnosed COVID-19 cases in the contributing population was estimated at 2.4 cases per 100,000 people. There are multiple limitations to this analysis because wastewater detection depends on factors other than incidence, such as sampling methods (e.g., frequency of sampling aliquots), which can influence the probability of capturing shed viral particles from an infected individual. Additionally, the estimate may vary based on site-specific clinical testing availability, wastewater sampling methods (e.g., composite sampling, freezing before processing) and laboratory processing (e.g., 4S extraction method, RT-qPCR). The estimation method for a WBE case detection limit presented here could benefit both COVID-19 WBE and other disease WBE by providing a systematic method to compare the case detection limits across studies.

## 5. CONCLUSION

- Wastewater N1 concentrations had a moderate correlation with geocoded clinical testing data (*τ*_unnormalized_=0.43). Normalization of SARS-CoV-2 N1 signal in wastewater to any biomarker did not improve the correlation with clinical testing data, likely because of the low variation in daily flow rates.
- Of the normalization biomarkers tested, crAssphage was the most promising due to low spatial and temporal variability and because crAssphage-normalized N1 had the strongest correlation with clinical testing data (*τ*_crAssphage_=0.38, *τ*_*Bacteroides*_=0.35, *τ*_PMMoV_=0.18).
- 18S rRNA was not suitable as a normalization biomarker due to its variability in sample concentrations, high degradation rate, and ubiquity as a laboratory contaminant.
- There was evidence of clinical undertesting at location N, which supports that wastewater testing could provide insights about COVID-19 trends in the population when clinical testing capabilities are limited.
- The COVID-19 outbreak at San Quentin prison (location Q) corresponded to a measured N1 concentration that was higher than any sewershed tested (4.89 x 10^3^ gene copies/mL).
- The wastewater-based epidemiology case detection limit using the 4S RNA extraction method on frozen samples was estimated to be 2.4 COVID-19 cases in 100,000 people.
- Lead time in wastewater over clinical testing varied from 0 to 3 weeks depending on the location, biomarker normalization, and testing turnaround time.
- Systematic approaches for determining a WBE case detection limit, biomarker normalization, and trendline smoothing were presented that can be applied across future WBE studies.

## Data Availability

Full dataset and associated code are available through GitHub.

https://zenodo.org/record/4730990#.YIxkrqlKgUo

## ACKNOWLEDGEMENTS

We thank Matt Metzger, Melissa Thornton, and the whole COVID-WEB team for support. We also thank our wastewater utility partners for facilitating and assisting with wastewater sampling and physicochemical measurements, including from East Bay Municipal Utility District (Florencio Gonzalez, Bill Chan, Gabriela Esparza, Paula Hansen, Kiley Kinnon, Nick Klumpp, Debra Mapp, Christine Pagtakhan, Daniel Siu, Dave Williams, Zach Wu, and Cheryl Yee), Central Contra Costa Sanitary District (Lori Schectel, Mary Lou Esparza, Blake Brown, Amanda Cauble), San Jose-Santa Clara Regional Wastewater Facility (RWF) Operations and Laboratory staff, and Central Marin Sanitation Agency. We thank the COVID-19 WBE Collaborative (https://www.covid19wbec.org/) community for discussions of methods and approaches. Additionally, we thank Robert Tjian, Sarah Stanley, Erik Van Dis, Thomas Graham, and Mira Chaplin. We gratefully acknowledge funding from The Catena Foundation as well as rapid response grants from the Center for Information Technology Research in the Interest of Society and the Innovative Genomics Institute at UC Berkeley to K.L.N.. H.D.G. and L.C.K were supported by the National Science Foundation (NSF) Graduate Research Fellowship [grant number DGE-1752814]. In addition, H.D.G was supported by the Berkeley Fellowship, and L.C.K. was supported by NSF INTERN through Re-Inventing the Nation’s Urban Water Infrastructure [grant number: 28139880-50542-C].

