## Supplementary Information for "Interpretation of temporal and spatial trends of SARS-CoV-2 RNA in San Francisco Bay Area wastewater"


Supplemental Methods 2

A. Wastewater composite sample collection, continued 2

B. Wastewater sample processing via the 4S method, continued 2

C. RT-qPCR plate setup and controls, continued 3

D. qPCR data processing 3

E. Clinical testing and population data, continued 4

Supplemental Figures and Tables 5

Author contributions 31

References 32

### Supplemental Methods

#### A. Wastewater composite sample collection, continued

Immediately after collection at location K, wastewater was mixed and aliquoted in 1-L bottles that were frozen at -20°C. These samples were transported together on ice and processed within 48 hours, and biological triplicates were taken from each bottle, where biological replicates refer to wastewater subsamples. All location K samples were processed in triplicate. Daily flow data throughout the study period were collected by the wastewater agency (Central Contra Costa Sanitary District) and used to calculate the mean flow rate.

Location E samples were stored onsite at -20°C, transported to the lab on ice, returned to storage at -20°C, thawed at 4°C, and then processed within 48 hours. Many of the samples from this site went through an additional freeze-thaw during transport despite being transported on ice. For location E, three sample replicates were processed for each date except 5/29/20 (2), 5/31/20 (2), 7/07/20 (2), 7/19/20 (2), 7/28/20 (2), 9/8/20 (1), and 9/14/20 (1). Daily flow data throughout the study period were collected by the wastewater agency (San Jose - Santa Clara Regional Wastewater Facility (SJSC-RWF)) and used to calculate the mean flow rate.

All other samples (locations S, A, N, and Q) were collected in 1-L bottles transported on ice within 48 hours of collection and then frozen at -80°C until processing. For location S, no samples had replication except 6/30 which was processed in triplicate. For location A, samples were processed in duplicate from 5/28/20 to 7/28/20 and with only one replicate from 8/4/20 to 9/9/20. For location N, no samples had biological replication. East Bay Municipal Utility District provided average sewershed flow rates for locations A, N, and S.

For location Q, no samples had replication except 7/1 which was processed in biological duplicate. For Central Marin Sanitation Agency only data from 6/1/20 to 7/13/20 were used in calculation of the average flow due to flow meter malfunction after this point.

#### B. Wastewater sample processing via the 4S method, continued

Briefly, sodium chloride was added to 40-50 mL of wastewater to a final concentration of 4 M, Ethylenediaminetetraacetic acid was added to a final concentration of 1 mM, and the solution was buffered using 10mM tris(hydroxymethyl)aminomethane to pH 7.2. Samples were heated to 70°C for 45 minutes and prefiltered with a 5-µm PVDF filter using syringe filtration. The filtrate was mixed with 40 mL of 70% ethanol and vacuum filtered through a silica column (Zymo III-P), and the column was washed using 5 mL of wash buffer 1 and 10 mL of wash buffer 2. Genetic material was eluted from the column by adding 200 µL ZymoPURE elution buffer and heating the column with elution buffer to 50°C for 10 minutes, then centrifuging the column to collect the flowthrough. The eluate was stored in multiple tubes to minimize freeze-thaw at -80°C until qPCR.

Each extraction batch contained a negative control of 40 mL of phosphate buffered saline (PBS) solution, and each sample or control was spiked with 20 µL of a free RNA control (SOC; stock solution of 1.33x$10^9$ gene copies/μL) and 50 μL of a surrogate virus lysis/extraction control from the same bottle of Bovilis Coronavirus Calf Vaccine (Merck Animal Health, Merck & Co. Inc., Kenilworth, NJ, USA) resuspended in 20 mL of PBS to monitor recovery with and without lysis across batches. Four representative samples were chosen from each location to assess SOC and BCoV recovery along with the batch PBS control, and Cts remained relatively consistent for SOC and varied considerably for BcoV. However, we saw no signs that an extraction procedure failed and considered all samples to pass this quality control screen.

#### C. RT-qPCR plate setup and controls, continued

To minimize qPCR contamination, sample processing and RT-qPCR plate assembly were performed in separate laboratories. Primers and probes were purchased as custom DNA oligos (Integrated DNA Technologies), except for the N1 assay (2019-nCoV CDC RUO Kit) and the Xeno assay (VetMAX™ Xeno™ Internal Positive Control - VIC™ Assay, ThermoFisher Scientific). Standard curves consisted of 10-fold serial dilutions of RNA standard from the same production batch of either synthetic RNA (Control 2- 102024, Twist Bioscience, San Francisco, CA) for the N1 assay, RNA from custom Ultramer RNA Oligonucleotides (Integrated DNA Technologies) for BCoV and PMMoV, geneBlocks DNA (Integrated DNA Technologies) for crAssphage, or RNA in-vitro transcribed from geneBlocks (Integrated DNA Technologies) with a HiScribe T7 Quick High Yield RNA Synthesis kit (New England Biolabs) for the *Bacteroides*, SOC, and 18S assays.

A subset of samples were run with no reverse transcription (No-RT) controls for *Bacteroides* rRNA and 18S rRNA assays to assess the relative contributions of RNA and DNA template to the target signal because both are expected to be present in samples. No-RT controls were conducted by heat-inactivating the reverse transcription enzyme in the Taqman Fast Virus One-Step Mastermix at 95°C for 5 minutes as per manufacturer's instructions before continuing with qPCR. Results were compared to the same samples with the RT step included. RNA was found to be multiple orders of magnitude greater than DNA in the samples tested, and results are given in **Table S7**. RNA and DNA yield were quantified via Qubit and found to be proportional (**Figure S1**).

#### D. qPCR data processing

Raw Cq values were imported into a custom pipeline in python (v3.6.9) with key modules including Pandas (v1.1.5) and NumPy (v1.19.5). First, raw Cq values that did not amplify or that amplified below the limit of detection were substituted with the Cq value corresponding to half the limit of detection (for N1) or half the bottom of the master standard curve (for all other assays) (**Table S5**) so that unamplified values could be considered during outlier analysis. The N1 limit of detection (LoD) was calculated by analyzing all the RNA standard curves from the study as well as four additional triplicate standard curves that extended down to 0.3 gc/μL (**Table S8**). The N1 LoD was set at 5 gc/rxn, at which point 67% of technical replicates were positive (**Table S8**). The number of true unamplified values was also determined prior to substitution. 24 samples were deemed below the N1 limit of detection (**Table S12**) and were set to half the detection limit. For normalization purposes only, all samples that were below the detection limit for the N1 assay were divided by the upper quartile value for that biomarker within each location instead of the measured value, such that when N1 values below the detection limit were normalized, all values were equal. Next, outlier testing was performed using a two-sided Grubbs Test (alpha= 0.05; scikit-learn v0.22.post1). Raw Cq values that did not pass scikit-learn Grubbs test were removed from further analysis. Next, Cq values were combined by calculating the average of the remaining values. Finally, the individual standard curve information was determined (**Table S4**) for validation after outlier assessment, but Cq values were converted to quantities (gene copies per reaction) using the master standard curves (**Table S5**). Individual standard curve efficiencies ranged from 83.2% to 97.8%, and $R^2$ ranged from 0.974 to 0.999 (**Table S4**). NTCs only amplified for the SOC assay and they amplified far outside the range of the standard curve (Cq of the NTCs= 38 and Cq of the bottom of the standard curve = 29; **Tables S4 and S5**). qPCR quantities were converted to gene copies per mL using the weight-based volume of the wastewater samples and the elution volume after the 4S extraction. For samples with biological replicates (**Table 1**), the geometric mean and standard deviation of the biological replicates were calculated (SciPy v1.4.1) and used to plot points and error bars respectively.

#### E. Clinical testing and population data, continued

For daily new cases from locations S, K, A, and N, values below 11 new cases per day were masked by public health departments to maintain confidentiality of the contributing population and substituted at 5.5 cases for further analysis (**Figures 2, 4 and S12**). For Location S, daily new case data (masked) and seven day moving averages were provided (unmasked because all values were >11). For location K, daily new case data (masked) were provided, and seven day moving averages of daily new cases were then calculated. Due to the low number of cases in locations A and N, most of the daily new case data were masked and are not shown. For location A, seven day moving averages (masked) were provided. For location N, fourteen day moving averages (masked) were provided. These moving averages of new COVID-19 cases per day were divided by the sewershed population (daily per capita cases) (**Table 1**). Population data were provided by East Bay Municipal Utility District for locations S, A, and N. Population data for location K was provided by the Contra Costa County Public Health Department.

For San Quentin Prison (location Q), COVID-19 clinical data were obtained from the California Department of Corrections and Rehabilitation open data portal (1) (**Table 1**). These data included TotalConfirmed, defined as "the cumulative number of patients with a positive COVID-19 result," which was divided by population to estimate new cases reported on each day. Population and prison capacity data were found in population reports (2). No data were masked by the California Department of Corrections and Rehabilitation for location Q, but instances of zero cases were substituted at 0.5 cases for comparison to masked data in statistical data analysis (**Figure 5**).

### Supplemental Figures and Tables

**Table S1:** Physicochemical wastewater parameters for each sampling location. Cells contain "Not available" when parameters were not measured by the corresponding utility. For location Q, the flow meter malfunctioned midway through the study, and data are only reported from before that date. Only mean flow rates, not daily, were available for locations S, N, and A.

<table>
<tr><th>Location</th><th>Mean flow rate (gal/person/day)</th><th>Summary statistic</th><th>Daily flow rate (MGD)</th><th>TSS (mg/L)</th><th>COD (mg/L)</th><th>BOD (mg/L)</th><th>cBOD (mg/L)</th></tr>
<tr><td rowspan="3">K</td><td rowspan="3">68.9</td><td>mean</td><td>33.3</td><td>262</td><td>586</td><td rowspan="3">Not available</td><td>208</td></tr>
<tr><td>cov</td><td>2%</td><td>8%</td><td>5%</td><td>12%</td></tr>
<tr><td>n</td><td>122</td><td>118</td><td>18</td><td>65</td></tr>
<tr><td rowspan="3">S, N, A</td><td rowspan="3">74.5, 71.9, 72.4</td><td>mean</td><td>35, 10, 6</td><td rowspan="3">Not available</td><td rowspan="3">Not available</td><td rowspan="3">Not available</td><td rowspan="3">Not available</td></tr>
<tr><td>cov</td><td>-</td></tr>
<tr><td>n</td><td>-</td></tr>
<tr><td rowspan="3">Q</td><td rowspan="3">127</td><td>mean</td><td>0.41</td><td>309</td><td rowspan="3">Not available</td><td>309</td><td rowspan="3">Not available</td></tr>
<tr><td>cov</td><td>4%</td><td>33%</td><td>58%</td></tr>
<tr><td>n</td><td>43</td><td>14</td><td>13</td></tr>
<tr><td rowspan="3">E</td><td rowspan="3">66.6</td><td>mean</td><td>100</td><td>289</td><td>590</td><td>275</td><td rowspan="3">Not available</td></tr>
<tr><td>cov</td><td>4%</td><td>7%</td><td>8%</td><td>10%</td></tr>
<tr><td>n</td><td>107</td><td>41</td><td>37</td><td>37</td></tr>
</table>

**Table S2** RT-qPCR reaction conditions for each assay. All reaction volumes were 20 µL.

| Reaction Component | Duplexed N1-Xeno (mM) | PMMoV (mM) | BCoV (mM) | 18S (mM) | SOC (mM) | CrAss-phage (mM) | Bacter-oides (mM) |
|---|---|---|---|---|---|---|---|
| TaqMan Fast Virus 1-Step Master Mix | 1x | 1x | 1x | 1x | 1x | 1x | 1x |
| Primer F | 0.5 | 0.4 | 0.9 | 0.5 | 0.5 | 0.5 | 0.5 |
| Primer R | 0.5 | 0.4 | 0.9 | 0.5 | 0.5 | 0.5 | 0.5 |
| Probe | 0.125 | 0.2 | 0.25 | 0.125 | 0.125 | 0.125 | 0.125 |
| Xeno Assay | Proprietary (0.8 µL/rxn) | - | - | - | - | - | - |
| Xeno RNA | 50 cp/µL | - | - | - | - | - | - |

**Table S3:** RT-qPCR thermocycling conditions for all assays

| Reaction Cycling Step | Temperature (°C) | Time (minutes: seconds) |
|---|---|---|
| UNG incubation | 25 | 2:00 |
| RT step | 50 | 15:00 |
| Polymerase activation | 95 | 2:00 |
| 45 cycles | 95 | 0:03 |
| | 55 | 0:30 |

**Table S4:** qPCR assay information for the SARS-CoV-2 nucleocapsid N gene (N1), the bovine coronavirus transmembrane protein gene (BCoV), the pepper mild mottle virus coat protein gene (PMMoV), human 18S ribosomal rRNA (18S), crAssphage 056 (crAssphage), *Bacteroides* 16S rRNA (*Bacteroides*)

| **Gene Target** (Reference) | **Type of Sequence** (length; accession) | **Sequence** (5’ -> 3’) |
|---|---|---|
| SARS-CoV-2 N1 (NA) | Forward primer | GACCCCAAAATCAGCGAAAT |
| | Reverse primer | TCTGGTTACTGCCAGTTGAATCTG |
| | Probe | FAM-ACCCCGCATTACGTTTGGTGGACC- ZEN/IBFQ |
| | Amplicon (72 bp; MN908947.3) | GACCCCAAAATCAGCGAAATGCACCCCGCATTACGTTTG GTGGACCCTCAGATTCAACTGGCAGTAACCAGA |
| VetMAX™ Xeno™ Internal Positive Contro (NA)l | Forward primer | Proprietary |
| | Reverse primer | Proprietary |
| | Probe | Proprietary |
| | Amplicon | Proprietary |
| PMMoV (3) | Forward primer | GAGTGGTTTGACCTTAACGTTTGA |
| | Reverse primer | TTGTCGGTTGCAATGCAAGT |
| | Probe | FAM-CCTACCGAAGCAAATG-ZEN/IBFQ |
| | Amplicon (68 bp; AB716964) | GAGTGGTTTGACCTTAACGTTTGAGCGGCCTACCGAAGC AAATGTCGCACTTGCATTGCAACCGACAA |
| BCoV (4) | Forward primer | CTGGAAGTTGGTGGAGTT |
| | Reverse primer | ATTATCGGCCTAACATACATC |
| | Probe | FAM-CCTTCATATCTATACACATCAAGTTGTT- ZEN/IBFQ |

| | | |
|---|---|---|
| | Amplicon (85 bp; AF39154) | CTGGAAGTTGGTGGAGTTTCAACCCAGAAACAAACAACT TGATGTGTATAGATATGAAGGGAAGGATGTATGTTAGGC CGATAAT |
| Human 18S rRNA (5) | Forward primer | GGTTCCTTTGGTCGCTCGCT |
| | Reverse primer | GGGCTGACCGGGTTGGTTTT |
| | Probe | FAM/AG AGC TAA T/ZEN/A CAT GCC GAC GGG C/IBFQ/ |
| | Amplicon (138bp; 6G18_2) | GGTTCCTTTGGTCGCTCGCTCCTCTCCTACTTGGATAACTG TGGTAATTCTAGAGCTAATACATGCCGACGGGCGCTGAC CCCCTTCGCGGGGGGGATGCGTGCATTTATCAGATCAAA ACCAACCCGGTCAGCCC |
| crAssphage (6) | Forward primer 056F1 | CAGAAGTACAAACTCCTAAAAAACGTAGAG |
| | Reverse primer 056R1 | GATGACCAATAAACAAGCCATTAGC |
| | Probe 056P1 | FAM-AATAACGATTTACGTGATGTAA-ZEN/IBFQ |
| | Amplicon (126bp; MT006214.1) | CAGAAGTACAAACTCCTAAAAAACGTAGAGGTAGAGGTA TTAATAACGATTTACGTGATGTAACTCGTAAAAAGTTTGA TGAACATACTGATTGTAATAAAGCTAATGGCTTGTTTATT GGTCATC |
| *Bacteroides* 16S rRNA (7) | Forward primer HF183 | ATCATGAGTTCACATGTCCG |
| | Reverse primer BacR287 | CTTCCTCTCAGAACCCCTATCC |
| | Probe | FAM/ATCGTTGAC/ZEN/TAGGTGGGCCGTTAC/IBFQ |
| | Amplicon (126 bp; MT464394.1) | ATCATGAGTTCACATGTCCGCATGATTAAAGGTATTTTCC GGTAGACGATGGGGATGCGTTCCATTAGATAGTAGGCGG GGTAACGGCCCACCTAGTCAACGATGGATAGGGGTTCTG AGAGGAAG |
| SOC (8) | Forward primer | CCACCAAAGTGGGCGATAAA |
| | Reverse primer | GGTGCCATTCGCCTCAATAA |
| | Probe | FAM/TGGCGGTGAGGAAGTTTGGAAAGA/ZEN/IBFQ |
| | Amplicon (89 bp, NA) | CCACCAAAGTGGGCGATAAAGGCAGCACCCGTTTATTTG GCGGTGAGGAAGTTTGGAAAGATAGCCCGATTATTGAGG CGAATGGCACC |

**Table S5**: All N1 assay RT-qPCR plate-specific standard curves after outlier assessment throughout the study.

| plate ID | linear dynamic range | slope | y-intercept | $R^2$ | PCR efficiency | Minimum Cq of NTC triplicates |
|---|---|---|---|---|---|---|
| 87 | 6 | -3.8 | 40.31 | 0.9928 | 0.832 | negative |
| 88 | 7 | -3.4 | 39.21 | 0.9989 | 0.967 | negative |
| 92 | 7 | -3.56 | 39.69 | 0.9921 | 0.91 | negative |
| 93 | 7 | -3.42 | 40.47 | 0.9839 | 0.962 | negative |
| 94 | 7 | -3.44 | 40.6 | 0.9968 | 0.954 | negative |
| 95 | 7 | -3.42 | 38.91 | 0.9976 | 0.962 | negative |
| 96 | 7 | -3.46 | 39.28 | 0.9992 | 0.945 | negative |
| 99 | 7 | -3.54 | 40.09 | 0.9954 | 0.915 | negative |
| 100 | 7 | -3.51 | 39.56 | 0.9944 | 0.927 | negative |
| 101 | 7 | -3.57 | 40.49 | 0.9964 | 0.905 | negative |
| 102 | 7 | -3.38 | 38.79 | 0.9919 | 0.978 | negative |
| 127 | 7 | -3.65 | 40.35 | 0.993 | 0.879 | negative |

**Table S6**: Master standard curve parameters (calculated after outlier assessment) and the values substituted for each assay for samples below the qPCR limit of detection

| Target | slope | y-intercept | substitution for BLoD samples (gc/rxn, Cq) | Min (gc/rxn, Cq) | Max (gc/rxn, Cq) | PCR efficiency | $R^2$ |
|---|---|---|---|---|---|---|---|
| N1 | -3.48 | 39.78 | 2.5, 38.39 | 5, 37.35 | 1E5, 22.4 | 0.94 | 0.986 |
| SOC | -3.52 | 42.02 | 5000, 29.01 | 10000, 27.95 | 1E10, 6.85 | 0.92 | 0.997 |
| BCoV | -3.83 | 47.27 | 50, 40.76 | 100, 39.61 | 1E8, 16.63 | 0.82 | 0.996 |
| PMMoV | -3.50 | 43.65 | 50, 37.71 | 100, 36.66 | 1E8, 15.67 | 0.93 | 0.995 |
| crAss-phage | -3.56 | 43.85 | 500, 34.24 | 1000, 33.17 | 1E9, 11.81 | 0.91 | 0.996 |
| Bacter-oides | -3.65 | 43.68 | 50, 37.47 | 100, 36.37 | 2E8, 13.36 | 0.88 | 0.985 |
| 18S | -3.76 | 43.51 | 50, 37.12 | 100, 35.99 | 1E8, 13.43 | 0.85 | 0.989 |

**Table S7:** Subset of samples tested with the reverse transcription step (RT) and without (no-RT controls) for the 18S and *Bacteroides* assays. Including the RT step should quantify both RNA and DNA, while the no-RT controls should only quantify DNA. RNA was orders of magnitude greater than DNA in these samples for both assays.

| Assay | Sample | RT-qPCR Concentration (cp/rxn) | qPCR (No-RT) Concentration (cp/rxn) | Concentration Ratio (RT / no-RT) |
|---|---|---|---|---|
| 18S | K_K_INF_061220_3 | 1.00E+05 | 6.90E+02 | 1.45E+02 |
| | K_K_INF_071020_3 | 2.00E+04 | 2.60E+02 | 7.69E+01 |
| | K_K_INF_080720_2 | 2.80E+05 | 8.50E+02 | 3.29E+02 |
| | K_K_INF_090420_2 | 1.10E+05 | 3.30E+02 | 3.33E+02 |
| *Bacteroides* | K_K_INF_061220_2 | 5.60E+06 | 5.10E+03 | 1.10E+03 |
| | K_K_INF_071020_2 | 2.10E+06 | 1.60E+03 | 1.31E+03 |
| | K_K_INF_080720_2 | 3.00E+06 | 2.70E+04 | 1.11E+02 |
| | K_K_INF_090420_2 | 4.40E+06 | 5.80E+04 | 7.59E+01 |

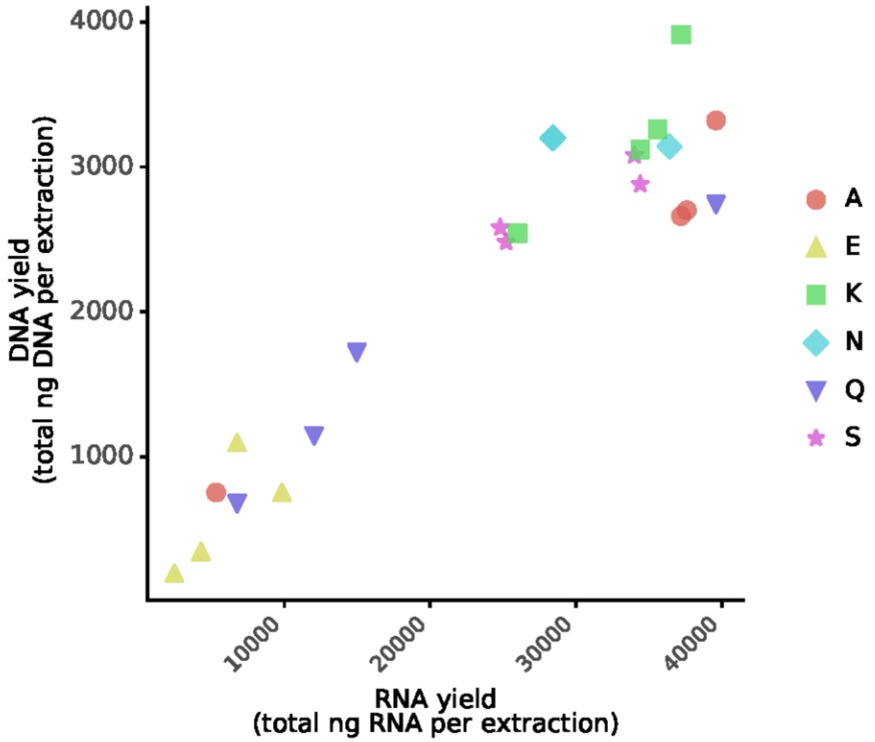


**Figure S1**: RNA yield and DNA yield from extracted wastewater samples using the 4S extraction method were proportional when assessed via Qubit.

**Table S8:** Evidence for the qPCR limit of detection for the N1 assay which was chosen as 5 gene copies per reaction. This table includes all N1 standard curves run throughout the study and four additional extended standard curves with quantities below 5 gene copies per reaction. All standard curves were processed in triplicate.

| standard curve Quantity (gene copies per reaction) | fraction of replicates positive | total number of replicates |
|---|---|---|
| **0.312** | **0.08** | **12** |
| **0.625** | **0.08** | **12** |
| **1.25** | **0.25** | **12** |
| **2.5** | **0.50** | **12** |
| **5** | **0.67** | **54** |
| **10** | **0.90** | **51** |
| **20** | **0.98** | **54** |
| **100** | **0.98** | **54** |
| **1000** | **1** | **54** |
| **10000** | **1** | **54** |
| **100000** | **0.98** | **54** |

**Table S9:** Inhibition testing for samples with Xeno dCt>2 relative to baseline in the pre-screen

| A | B | C | D | E | F | G | H | I | J | K | L |
|---|---|---|---|---|---|---|---|---|---|---|---|
| **Sample Location** Date | **Dilution factor** | **N1 Mean Quantity Adjusted for Dilution** | **N1 mean Ct** | **Unamplified N1 Replicates (%)** | **dCt** *N1 relative to 1x* | **[Expected dCt] – [Actual dCt]** *N1 relative to 1x* | **dCt** *N1 relative to previous* | **[Expected dCt] – [Actual dCt]** *N1 relative to previous* | **Inhibited** yes if subsequent dilution in column I > 1 | **Initial xeno dCt** *1x relative to baseline* | **Xeno dCt** *Relative to baseline* |
| **Q 09/15/20** | 1* | - | - | 100% | - | - | - | - | - | 2.6 | 1.62 |
| | 2 | - | - | 100% | - | - | - | - | - | | 1.38 |
| | 5 | - | - | 100% | - | - | - | - | - | | 0.76 |
| | 10 | - | - | 100% | - | - | - | - | - | | 0.87 |
| **A 09/09/20** | 1 | 5.32 | 36.85 | 0% | 0.00 | 0.00 | - | - | yes | 2.00 | 2.58 |
| | 2 | 12.87 | 36.79 | 33% | 0.58 | 0.42 | -0.06 | 1.06 | yes | | 1.77 |
| | 5 | 26.34 | 36.96 | 33% | 0.75 | 1.55 | 0.17 | 1.13 | yes | | 0.87 |
| | 10 | 81.24 | 36.22 | 0% | -0.34 | 3.64 | -0.74 | 1.74 | - | | 0.48 |
| **A 09/09/20 rerun** | 1* | 6.89 | 35.57 | 0% | 0.00 | 0.00 | - | - | no | | 2.79 |
| | 2 | 6.40 | 36.62 | 0% | 1.05 | -0.05 | 1.05 | 0.05 | no | | 2.16 |
| | 4 | 10.73 | 37.10 | 0% | 1.53 | 0.47 | 0.48 | 0.82 | no | | 1.63 |
| | 8 | 19.54 | 37.29 | 0% | 1.72 | 1.28 | 0.19 | 0.81 | - | | 0.42 |
| **N 08/11/20** | 1 | 32.87 | 33.89 | 0% | 0.00 | 0.00 | - | - | yes | 3.8 | 1.52 |
| | 2* | 111.73 | 33.18 | 0% | -0.71 | 1.71 | -0.71 | 1.71 | no | | 1.98 |
| | 5 | 71.60 | 35.17 | 0% | 1.28 | 1.02 | 1.99 | -0.69 | no | | 0.47 |
| | 10 | 47.18 | 36.99 | 33% | 3.10 | 0.20 | 1.82 | -0.82 | - | | 0.73 |
| **N 09/01/20** | 1 | 5.67 | 37.02 | 0% | 0.00 | 0.00 | - | - | yes | 2.1 | 1.64 |
| | 2* | 16.51 | 36.25 | 0% | -0.77 | 1.77 | -0.77 | 1.77 | - | | 0.91 |
| | 5 | - | - | 66% | - | - | - | - | - | | 0.85 |
| | 10 | - | - | 100% | - | - | - | - | - | | 0.55 |
| **S 06/02/20** | 1* | 26.29 | 34.53 | 0% | 0.00 | 0.00 | - | - | no | 3.4 | 2.90 |
| | 2 | 32.45 | 35.26 | 33% | 0.72 | 0.28 | 0.72 | 0.28 | no | | 1.89 |
| | 5 | 38.63 | 36.34 | 33% | 1.81 | 0.49 | 1.08 | 0.22 | - | | 0.52 |
| | 10 | - | - | 66% | - | - | - | - | - | | 0.43 |

| | | | | | | | | | | | |
|---|---|---|---|---|---|---|---|---|---|---|---|
| **S 06/09/20** | 1* | 15.01 | 35.57 | 0% | 0.00 | 0.00 | - | - | no | 2.8 | 0.49 |
| | 2 | 31.46 | 35.78 | 0% | 0.21 | 0.79 | 0.21 | 0.79 | no | | 0.34 |
| | 5 | 18.23 | 37.42 | 33% | 1.85 | 0.45 | 1.63 | -0.33 | - | | 0.43 |
| | 10 | - | - | 66% | - | - | - | - | - | | 0.53 |
| **S 07/14/20** | 1 | 42.68 | 33.56 | 0% | 0.00 | 0.00 | - | - | no | 3.1 | 1.50 |
| | 2 | 46.44 | 34.47 | 0% | 0.91 | 0.09 | 0.91 | 0.09 | no | | 1.04 |
| | 5 | 43.04 | 35.98 | 0% | 2.42 | -0.12 | 1.51 | -0.21 | no | | 0.88 |
| | 10 | 59.56 | 36.71 | 33% | 3.15 | 0.15 | 0.73 | 0.27 | - | | 0.36 |
| **S 07/14/20 rerun** | 1* | 63.43 | 33.14 | 0% | 0.00 | 0.00 | - | - | no | | 2.20 |
| | 2 | 70.92 | 34.01 | 0% | 0.87 | 0.13 | 0.87 | 0.13 | no | | 1.46 |
| | 5 | 91.68 | 35.59 | 0% | 2.45 | -0.15 | 1.58 | -0.28 | no | | 1.01 |
| | 10 | 105.94 | 35.89 | 0% | 2.75 | 0.55 | 0.30 | 0.70 | - | | 0.55 |
| **E 06/07/20** | 1* | - | - | 100% | - | - | - | - | - | 2.02 | 1.12 |
| | 2 | - | - | 100% | - | - | - | - | - | | 0.91 |
| | 5 | - | - | 100% | - | - | - | - | - | | 0.86 |
| | 10 | - | - | 100% | - | - | - | - | - | | 0.71 |
| **E 09/08/20** | 1 | 4.59 | 37.95 | 0% | 0.00 | 0.00 | - | - | yes | 2.2 | 0.88 |
| | 2* | 20.04 | 37.02 | 0% | -0.92 | 1.92 | -0.92 | 1.92 | - | | 0.85 |
| | 5 | - | - | 66% | - | - | - | - | - | | 0.31 |
| | 10 | - | - | 100% | - | - | - | - | - | | 0.54 |
| **E 09/14/20** | 1* | 7.16 | 37.27 | 33% | 0.00 | 0.00 | | - | no | 3.7 | 2.58 |
| | 2 | 11.16 | 37.75 | 33% | 0.48 | 0.52 | 0.48 | 0.52 | - | | 1.10 |
| | 5 | - | - | 50% | - | - | - | - | - | | 0.76 |
| | 10 | - | - | 100% | - | - | - | - | - | | 0.35 |

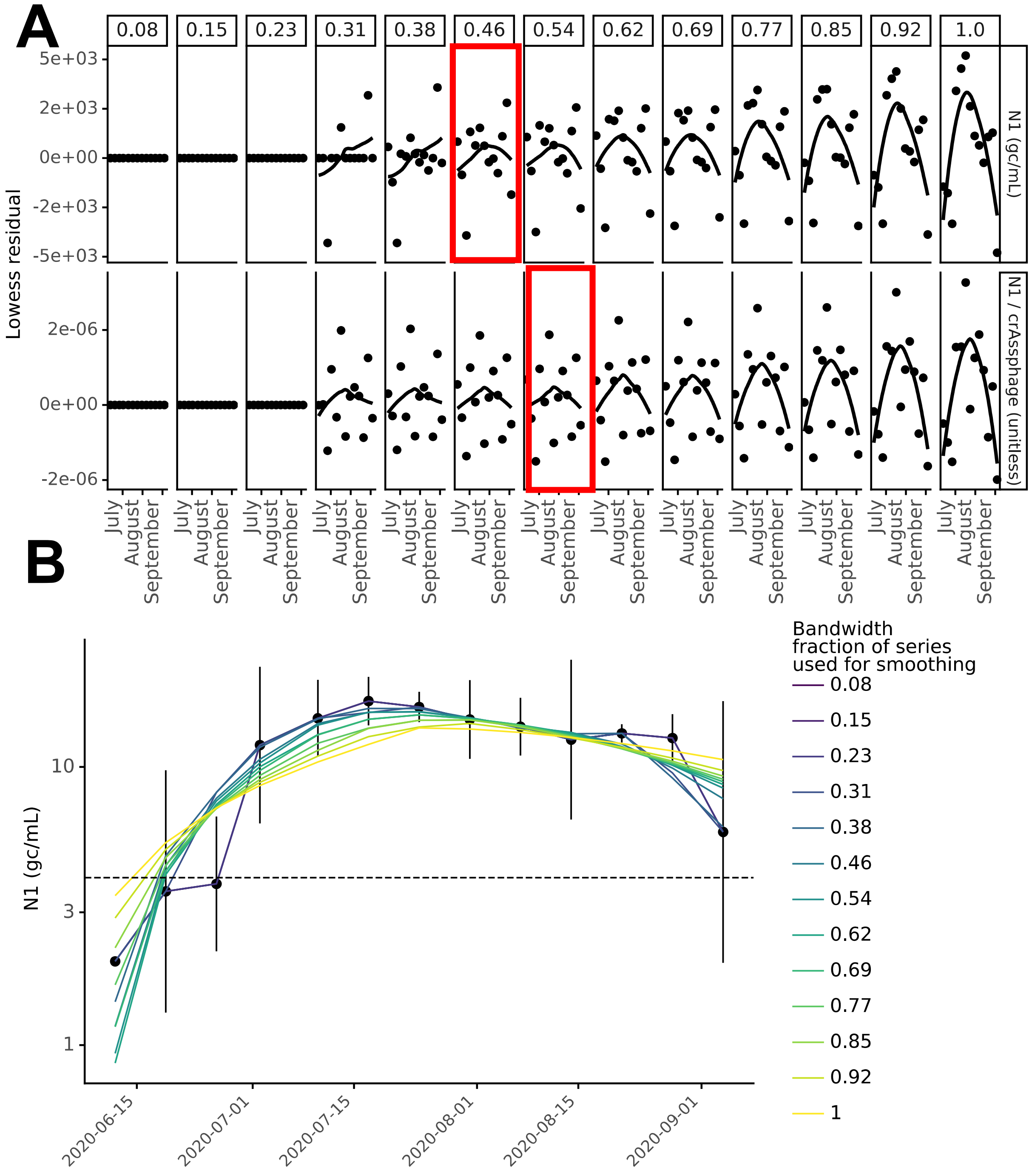


**Figure S2: Lowess bandwidth parameter selection process for Location K**

(A) Residual plots for Lowess bandwidth parameter (α; column labels) determination for location K where the bandwidth parameter increases from inclusion of 1 data point (far left) to inclusion of all data points (far right) in each local regression for unnormalized N1 (top) and crAssphage-normalized N1 (bottom). The value of α that minimized the residual was selected (red boxes). (B) Visualization of how bandwidth parameter affected the Lowess trendline for location K.

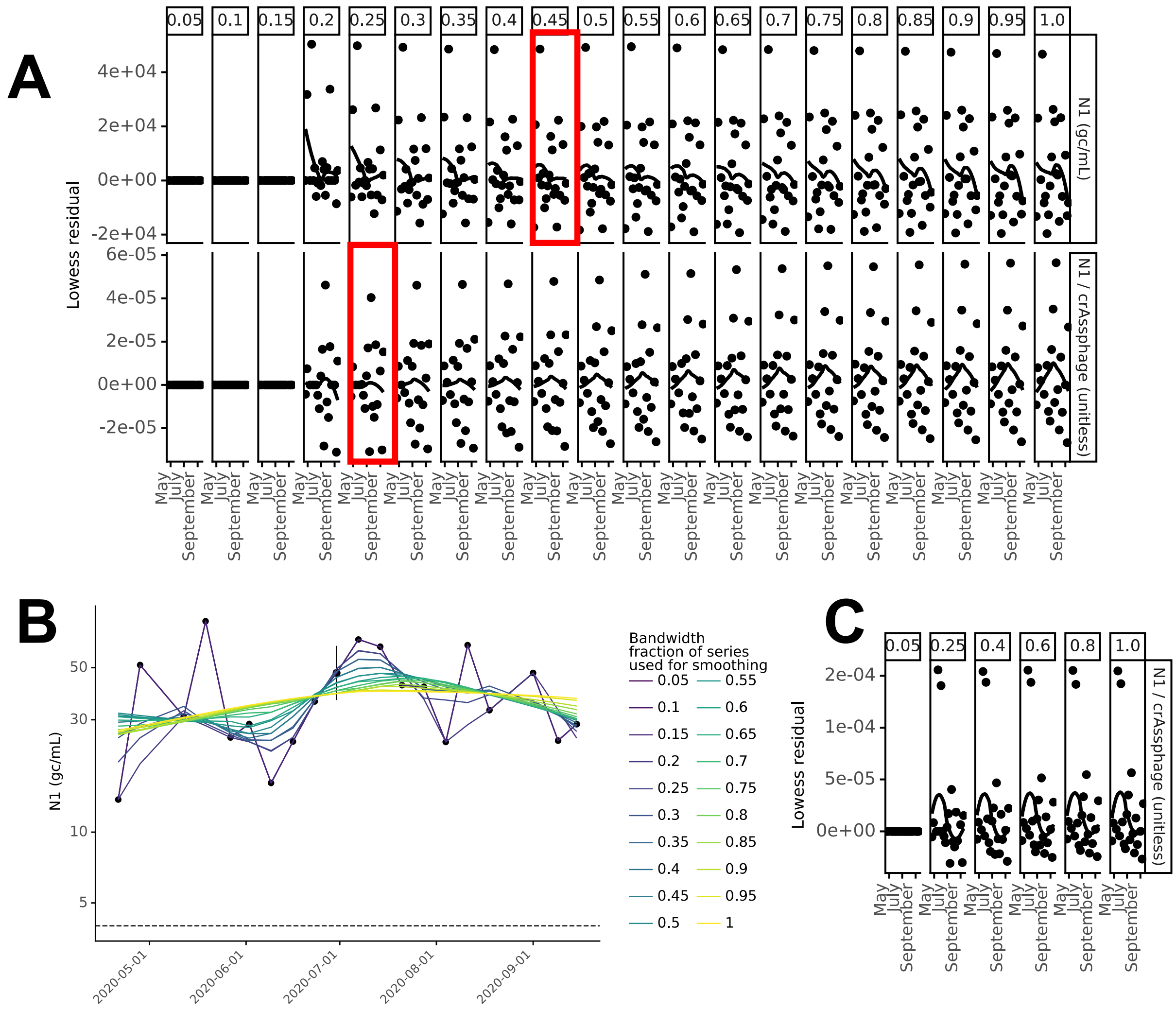


**Figure S3: Lowess bandwidth parameter selection process for Location S**
(A) Residual plots for Lowess bandwidth parameter (α; column labels) determination for location S where the bandwidth parameter increases from inclusion of 1 data point (far left) to inclusion of all data points (far right) in each local regression for unnormalized N1 (top) and crAssphage-normalized N1 (bottom). The value of α that minimized the residual was selected (red boxes). (B) Visualization of how bandwidth parameter affected the Lowess trendline for location S. (C) The Lowess residual of two points for crAssphage-normalized N1 obscured trends and were removed for selection of the bandwidth parameter in part A.

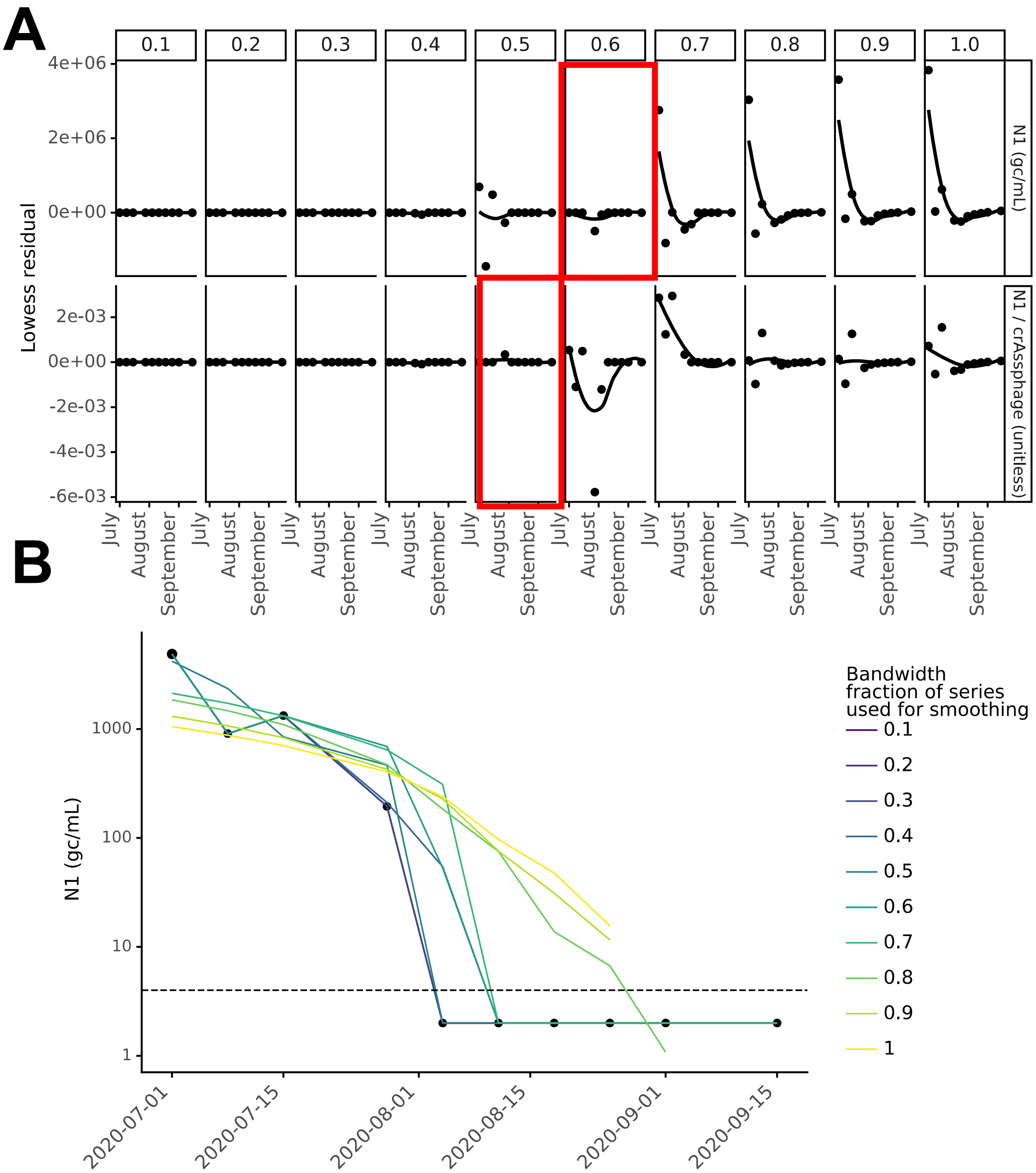


**Figure S4: Lowess bandwidth parameter selection process for Location Q**

(A) Residual plots for Lowess bandwidth parameter (α; column labels) determination for location Q where the bandwidth parameter increases from inclusion of 1 data point (far left) to inclusion of all data points (far right) in each local regression for unnormalized N1 (top) and crAssphage-normalized N1 (bottom). The value of α that minimized the residual was selected (red boxes). (B) Visualization of how bandwidth parameter affected the Lowess trendline for location Q.

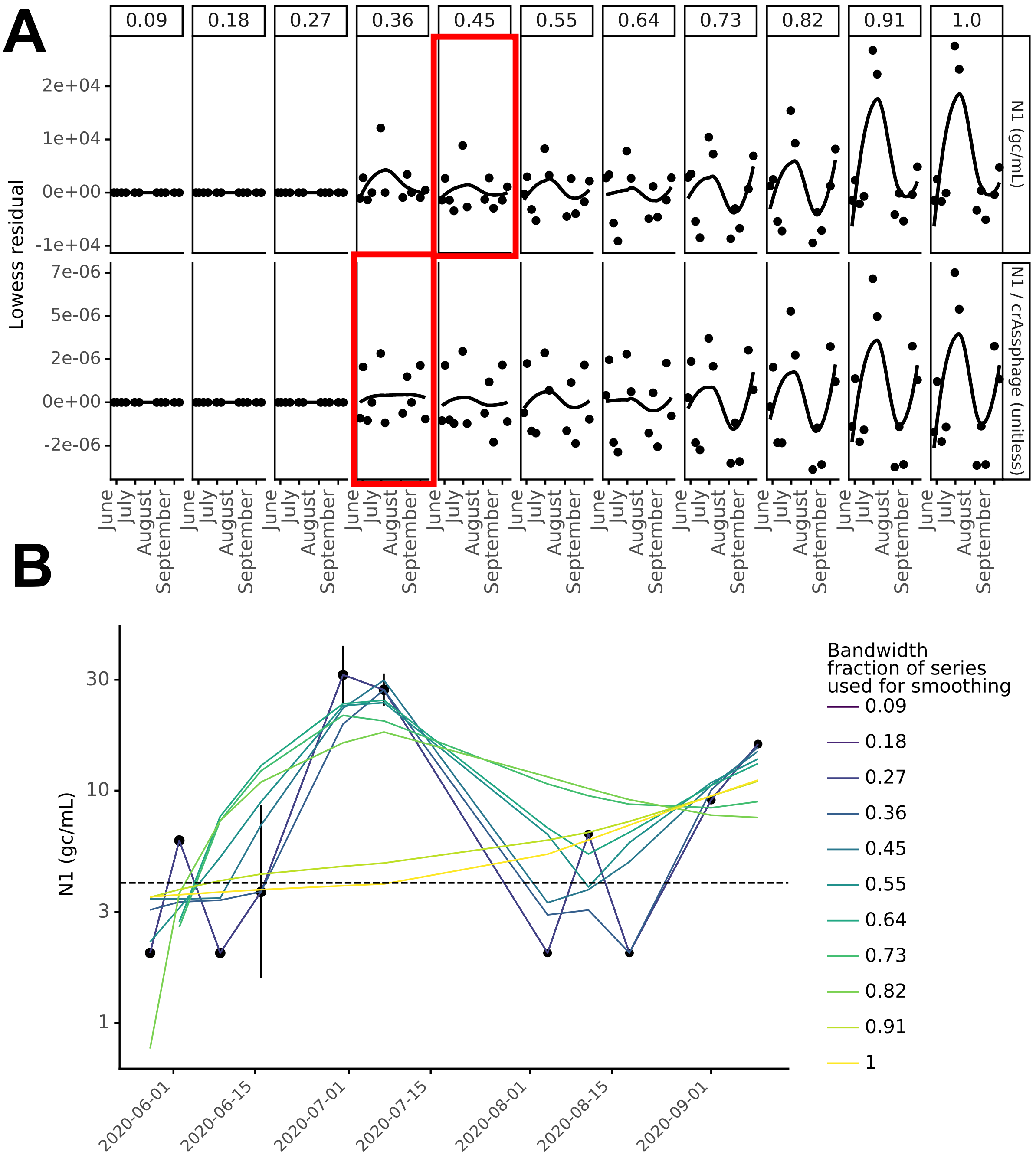


**Figure S5: Lowess bandwidth parameter selection process for Location A**

(A) Residual plots for Lowess bandwidth parameter (α; column labels) determination for location A where the bandwidth parameter increases from inclusion of 1 data point (far left) to inclusion of all data points (far right) in each local regression for unnormalized N1 (top) and crAssphage-normalized N1 (bottom). The value of α that minimized the residual was selected (red boxes). (B) Visualization of how bandwidth parameter affected the Lowess trendline for location A.

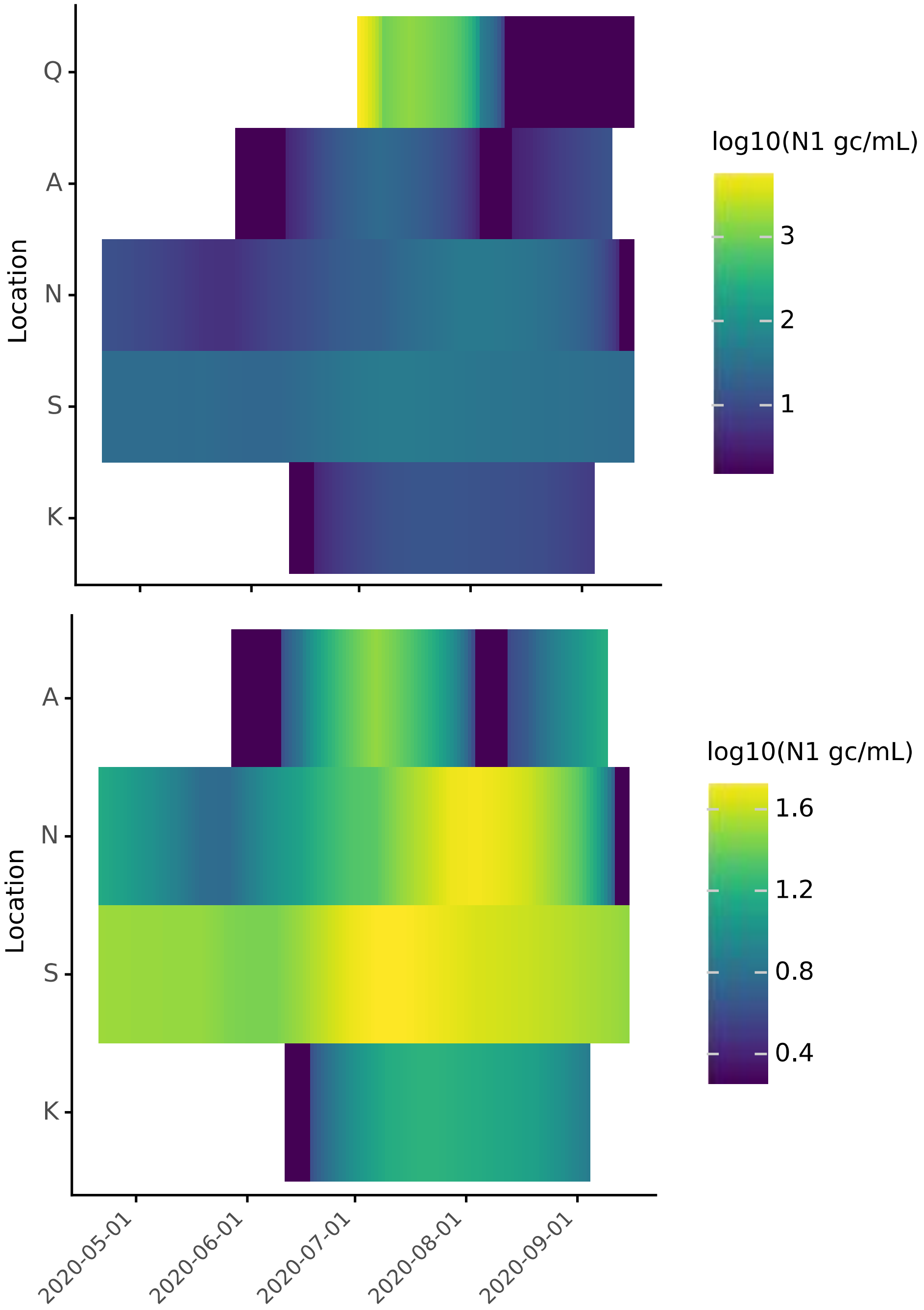


**Figure S6**: heatmap visualization of most optimal Lowess trendlines for SARS-CoV-2 N1 signal in wastewater by location **(top)** all locations **(bottom)** without location Q

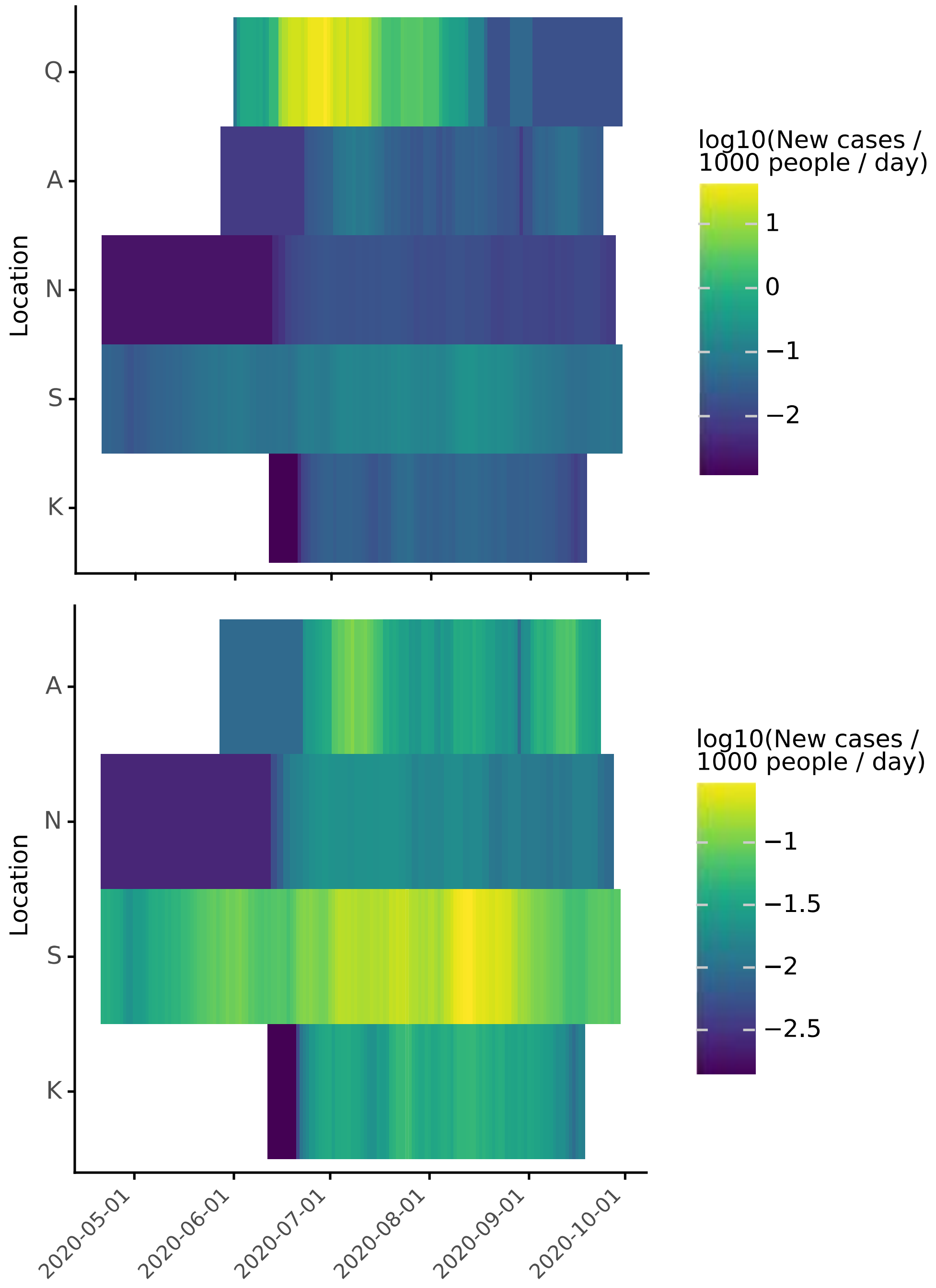


**Figure S7**: heatmap visualization of seven day moving average of COVID-19 daily per capita new cases by location **(top)** all locations **(bottom)** without location Q

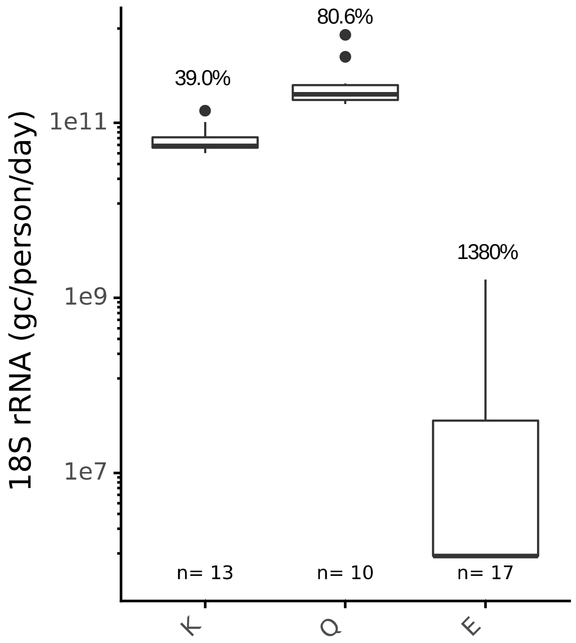


**Figure S8:** 18S rRNA flow-scaled values across K, E, and Q sampling locations with geometric coefficients of variation shown above the boxes. Samples that did not amplify or amplified below the detection limit were assigned a value half that of the assay detection limit. Because of the inconsistent detection and high variability in quantified samples, 18S was not measured in the remaining sampling locations (A, N, and S).

**Table S10:** Summary statistics of the normalization biomarkers tested in this study juxtaposed with ranges in literature values in raw wastewater. CrAssphage and PMMoV concentrations were consistent with past reported values in the literature, but to our knowledge, no prior raw wastewater values for *Bacteroides* rRNA (HF183) and 18S rRNA have been reported in the literature, although Cqs have been documented in raw wastewater and concentrations have been documented in other water types. We also measured HF183 rDNA (no RT step) in four samples and found concentrations to be 2.1-3.1 log lower than RNA (**Table S7**).

| Target | This study: Raw wastewater concentration geometric mean (gstd) | Literature: Range in raw wastewater concentration | Literature: Represented geographic range | Literature: References |
|---|---|---|---|---|
| crAssphage CPQ_056 and CPQ_064 | $1.21*10^{9}$ gc/L (1.985) | $6.92*10^{4}$- $2*10^{10}$ gc/L | United States, Australia, Chile, Thailand, Italy, Louisiana | (6,9–17) |
| PMMoV | $1.08*10^{8}$ gc/L (2.04) | $3.9*10^{4}$- $2.16*10^{10}$ gc/L | Australia, Florida, Arizona, Germany, Vietnam,Egypt | (9,12,15,18–27) |
| *Bacteroides* HF183 rRNA | $1.93*10^{10}$ gc/L (3.53) | - | - | - |
| *Bacteroides* HF183 rRNA gene | $8.48*10^{6}$ gc/L | $7*10^{7}$ - 3.1e*$10^{10}$ gc/L | United States, Chile, Belgium, Australia | (10,14,28,29) |
| Human 18S rRNA | $1.79*10^{7}$ gc/L (49.6) | - | - | - |

***Bacteroides* rRNA gene (DNA) was only measured in four samples as no reverse transcription controls

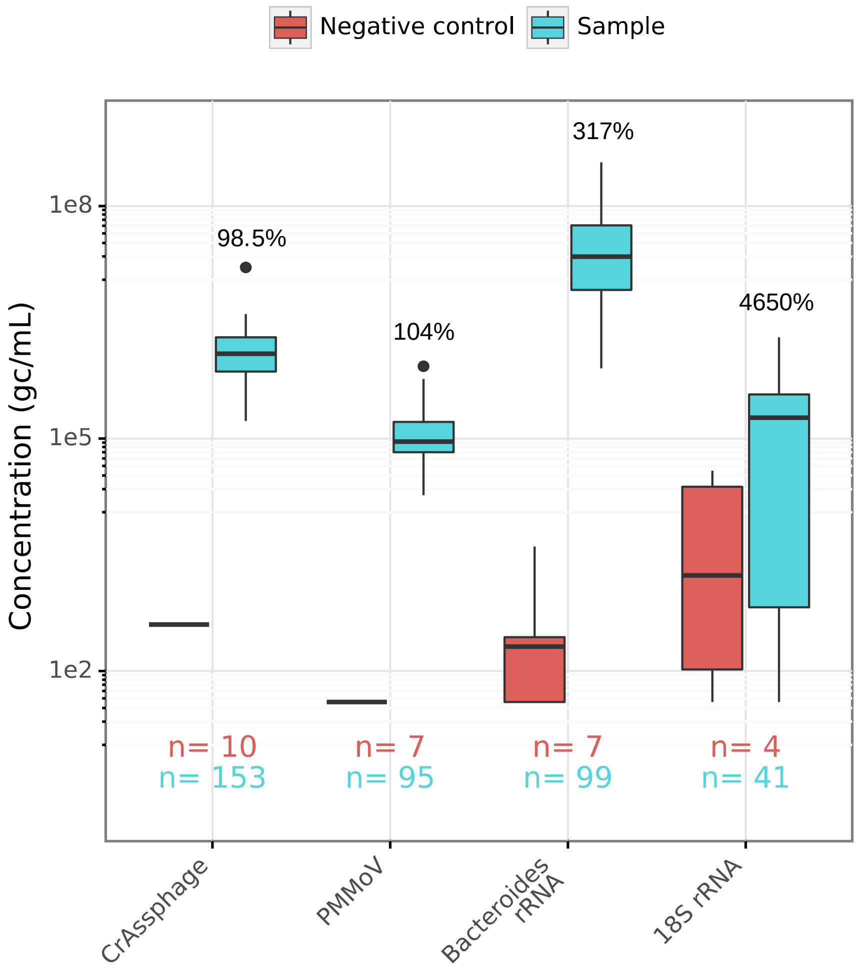


**Figure S9.** This figure illustrates the variability in the concentrations of normalization biomarkers across locations E, K, Q, A, N, and S. All biological replicates processed from all locations are included. Based on geometric coefficients of variation (shown above sample boxes), crAssphage and PMMoV display less variation across all the points in the study than do 18S and *Bacteroides*. Additionally, target concentrations in the extraction negative controls are displayed for comparison. When samples or negative controls were undetected via qPCR, the value is plotted as half of the limit of detection for the assay. No amplification of PMMoV or crAssphage occurred in extraction negative controls. Three out of four negative extraction controls amplified above the detection limit for the *Bacteroides* assay, but the concentrations are significantly different from the sample concentrations. Three of the four extraction negative controls were quantifiable via the 18S rRNA assay, and the concentrations of the samples and controls are not statistically different (p= 0.15).

**Table S11:** Unnormalized and normalized N1 correlations to clinical case data assessed via Kendall's tau-b. Datasets include locations K, S, N, A, and Q.

| | All data | | | Without BLoD | | |
|---|---|---|---|---|---|---|
| | **Correlation to case data** | **p value** | **n** | **Correlation to case data** | **p value** | **n** |
| **Unnormalized N1** | 0.43 | 1.85E-07 | 72 | 0.42 | 2.1E-05 | 49 |
| **N1 / crAssphage** | 0.38 | 2.48E-06 | 72 | 0.30 | 2.8E-03 | 49 |
| **N1 / PMMoV** | 0.18 | 3.14E-02 | 68 | 0.04 | 6.6E-01 | 47 |
| **N1 / *Bacteroides*** | 0.35 | 1.70E-05 | 72 | 0.31 | 1.3E-03 | 51 |

**Table S12:** By location, total number of biological replicates and detectable portion compared to the percent below the N1 limit of detection

| Location | Total number of biological replicates | Number of detectable biological replicates | Percent of biological replicates below the limit of detection |
|---|---|---|---|
| A | 17 | 10 | 41.2 |
| K | 39 | 32 | 17.9 |
| N | 18 | 14 | 22.2 |
| Q | 11 | 5 | 54.5 |
| S | 22 | 22 | 0 |
| total | 107 | 83 | 22.4 |

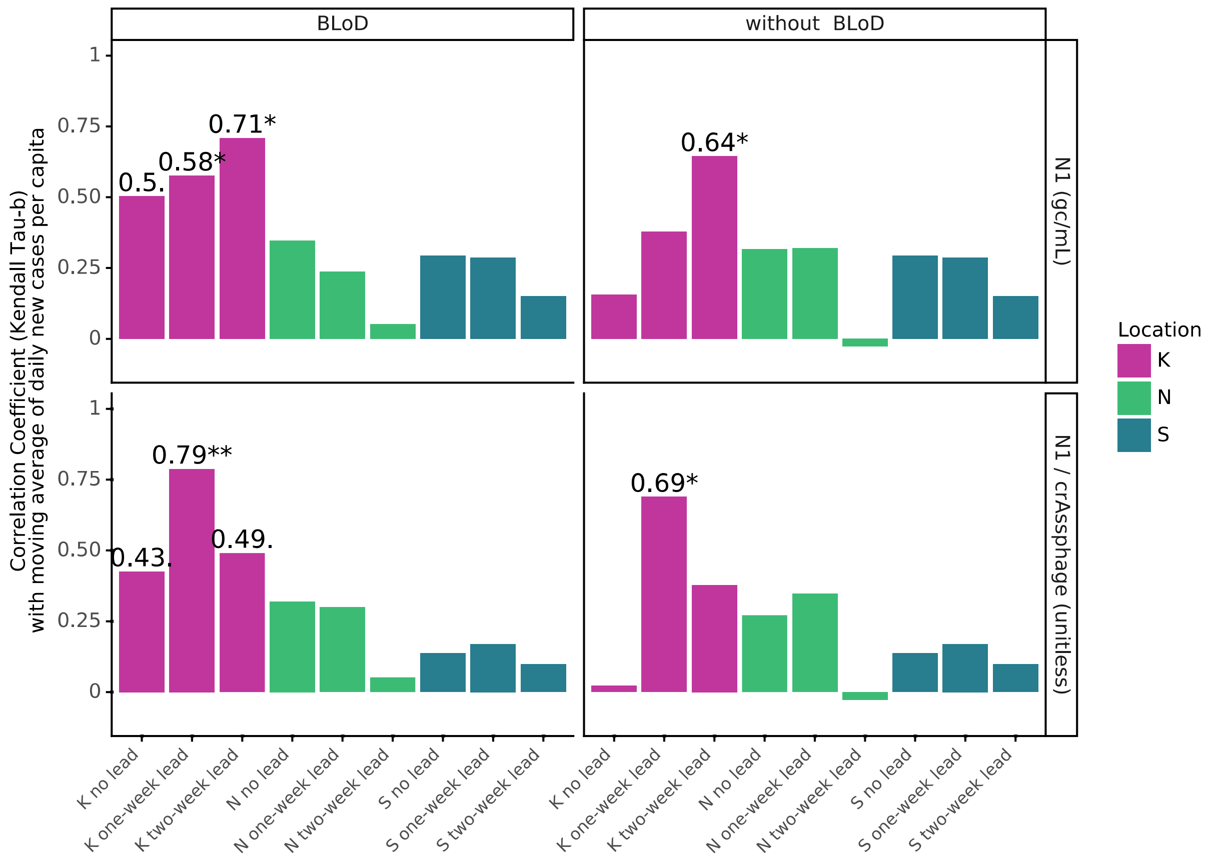


**Figure S10**: Kendall's Tau-b by location for comparisons between wastewater SARS-CoV-2 N1 signal (associated with the sample collection date) and COVID-19 cases (associated with the result date) both with and without samples below the N1 limit of detection (BLoD) included (column facets). Wastewater SARS-CoV-2 data (row facets; unnormalized N1 (gc/mL) and crAssphage-normalized N1 (unitless)) were compared to the seven-day moving average of geocoded COVID-19 new cases. The analysis was completed with wastewater date aligned with clinical date (no lead) as well as with one- and two-week lead times (wastewater leads clinical testing data by one or two weeks). Significance is indicated as <0.05 . , <0.01 *, and <0.001**. Data from location K are repeated from **Figure S14** for location comparison.

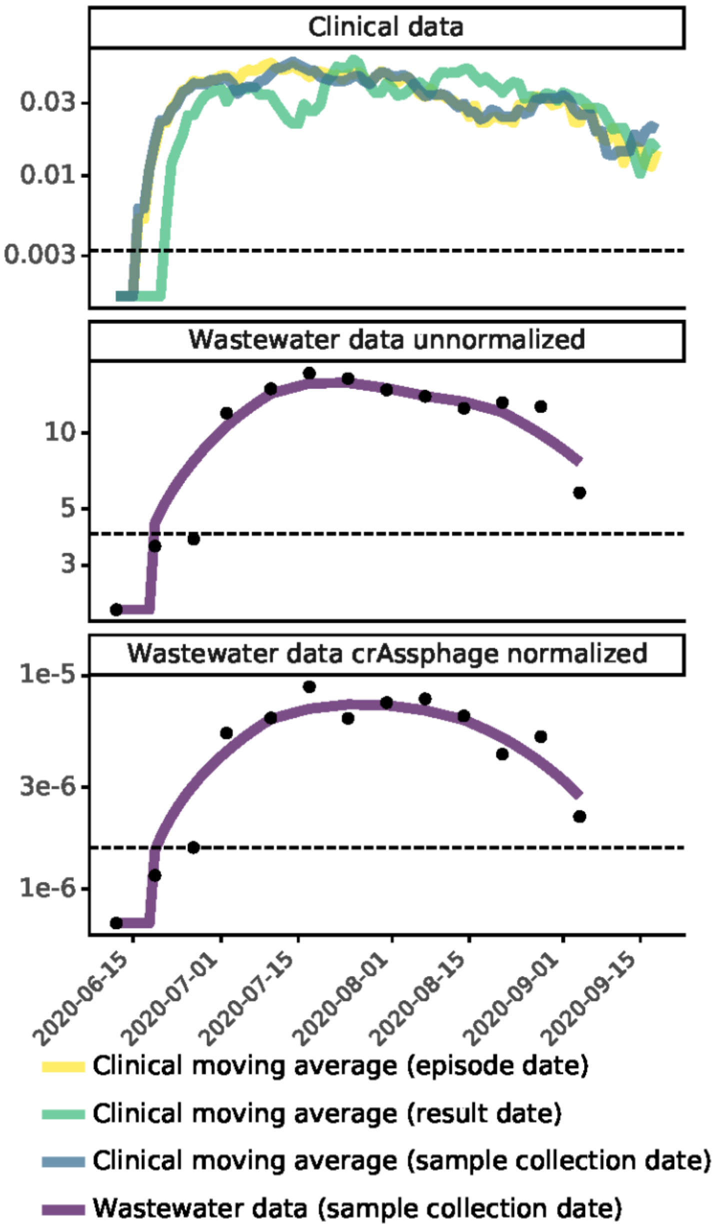


**Figure S11**: Comparison of geocoded COVID-19 clinical testing results (**top**) to wastewater SARS-CoV-2 N1 signal (**middle**) and crAssphage-normalized signal (**bottom**) at location K from June to September 2020. COVID-19 clinical testing results are the seven-day moving averages of daily per capita cases with data aligned by episode date (yellow line), result date (green line), or specimen collection date (blue line), with semi-transparency to visualize overlapping sections. Wastewater SARS-CoV-2 N1 signal is aligned by specimen collection date (lines are the most optimal Lowess trendlines).

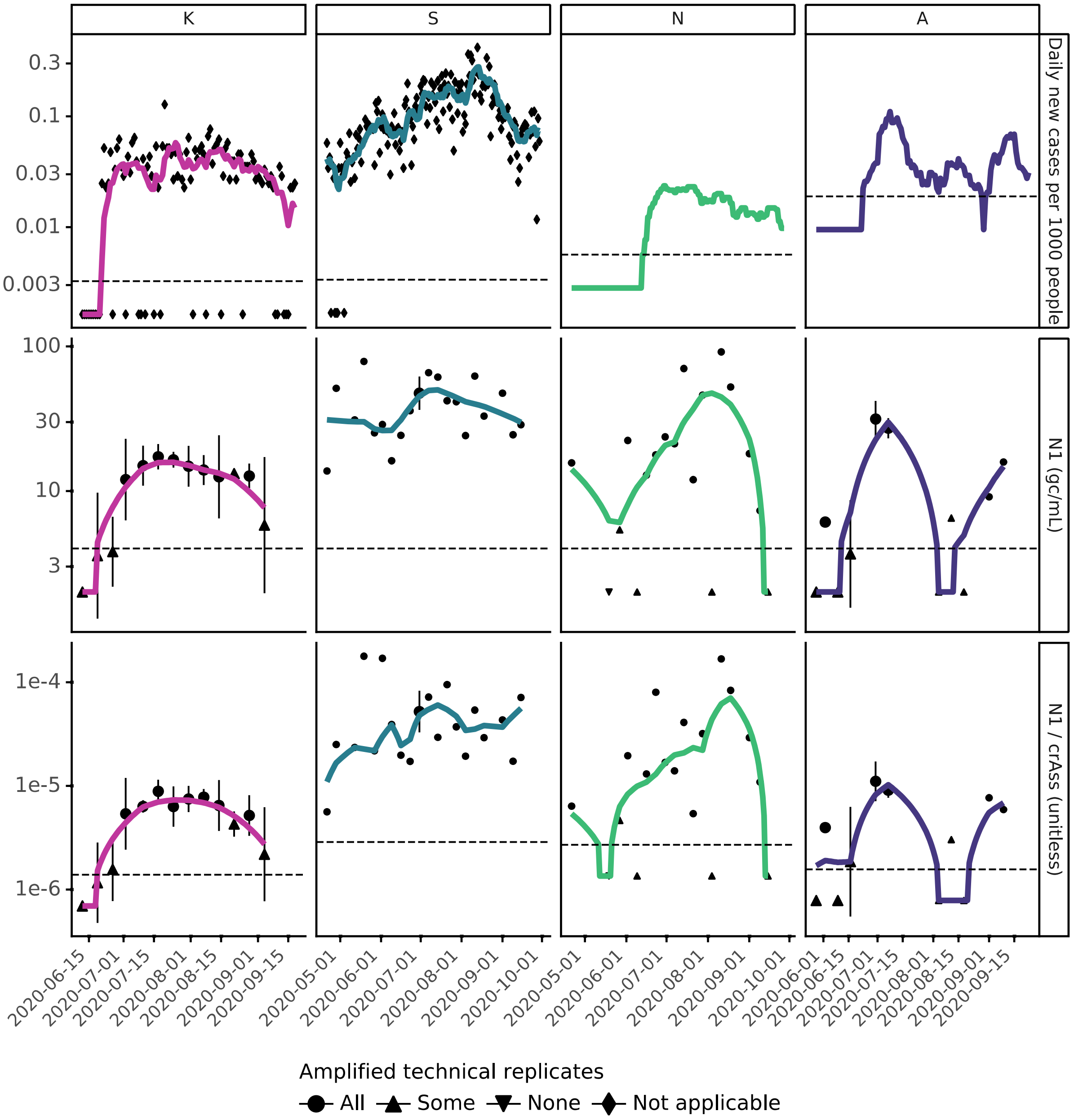


**Figure S12:** For locations serving more than 80,000 people: (**top**) SARS-CoV-2 N1 signal in wastewater, where symbols indicate the amount of technical replicates that amplified during qPCR, horizontal dashed line indicates the limit of detection, and solid lines represent the most optimal Lowess trendline (**Figures S2-5**). (**middle**) SARS-CoV-2 N1 signal normalized to crAssphage signal in wastewater, where symbols indicate the amount of technical replicates that amplified during qPCR, horizontal dashed line indicates the limit of detection for N1 divided by the upper quartile for crAssphage for each location, and solid lines represent the most optimal Lowess trendlines (**Figures S2-5**). (**bottom**) Daily COVID-19 clinical testing results from people within each sewershed normalized to the population in each sewershed, where symbols are plotted at the daily new cases per 1000 people per day, the trendline represents the seven-day moving average of these data, and the horizontal dashed line indicates 1 case in 1000 people for each location.

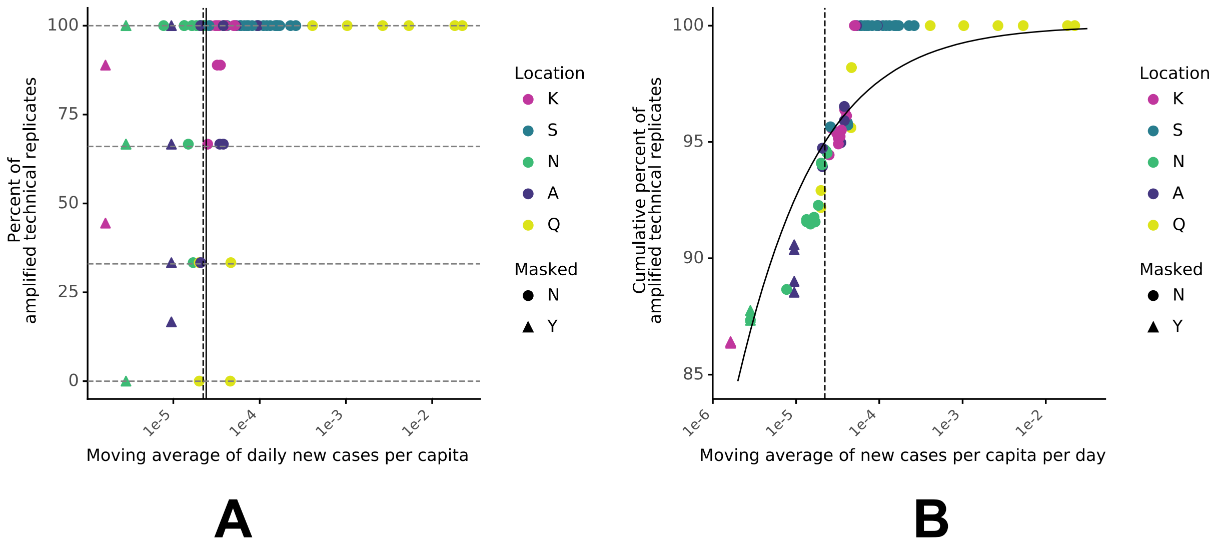


**Figure S13:** (**A**) The percent of amplified technical replicates for each value of the moving average of daily per capita cases (x-axis). Each biological replicate had three technical replicates, so the horizontal, dashed lines at 0%, 33%, 66%, and 100% show the range of values associated with each biological replicate that was associated with a unique x value. One or more biological replicates were associated with each moving average case value. The solid vertical line represents the estimated WBE clinical detection limit determined without masked data (2.4 cases in 100,000 people; **Figure 6**). (**B**) The cumulative percentage of amplified wastewater technical replicates was calculated by ranking the moving averages of daily per capita cases (including masked clinical case values; x-axis) from highest to lowest and calculating the fraction of qPCR replicates that amplified cumulatively (y-axis) for each value of x (same methodology as in **Figure 6**). In both plots, the dashed line represents the daily new cases per capita value above which 95% of wastewater technical replicates amplified (including masked values; 2.2 cases in 100,000 people), and shapes denote which daily per capita case values were masked.

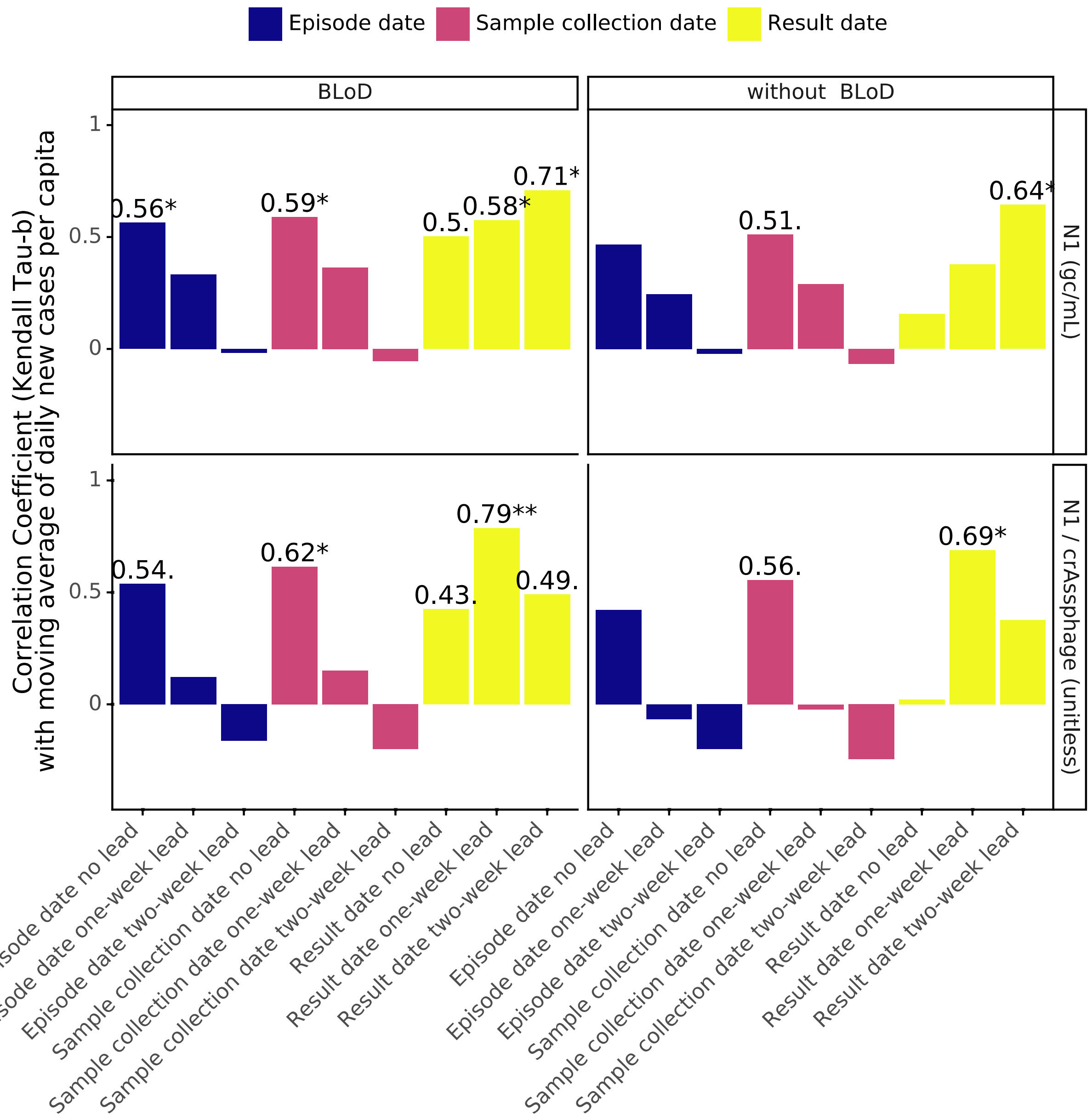


**Figure S14**: Kendall's Tau-b at location K for comparisons between wastewater SARS-CoV-2 N1 signal (associated with the sample collection date) and COVID-19 new cases (date association varies as indicated by color: episode date, specimen collection date, and result date) both with and without samples below the N1 limit of detection (BLoD) included (column facets). Wastewater SARS-CoV-2 data (row facets; unnormalized N1 (gc/mL) and crAssphage-normalized N1 (unitless)) were compared to the seven-day moving average of geocoded COVID-19 new cases. The analysis was completed with wastewater date aligned with clinical date (no lead) as well as with one- and two-week lead times (wastewater leads clinical testing data by one or two weeks). Significance is indicated as <0.05 . , <0.01 *, and <0.001**. Result date data are repeated from **Figure S10** for corresponding date comparison.

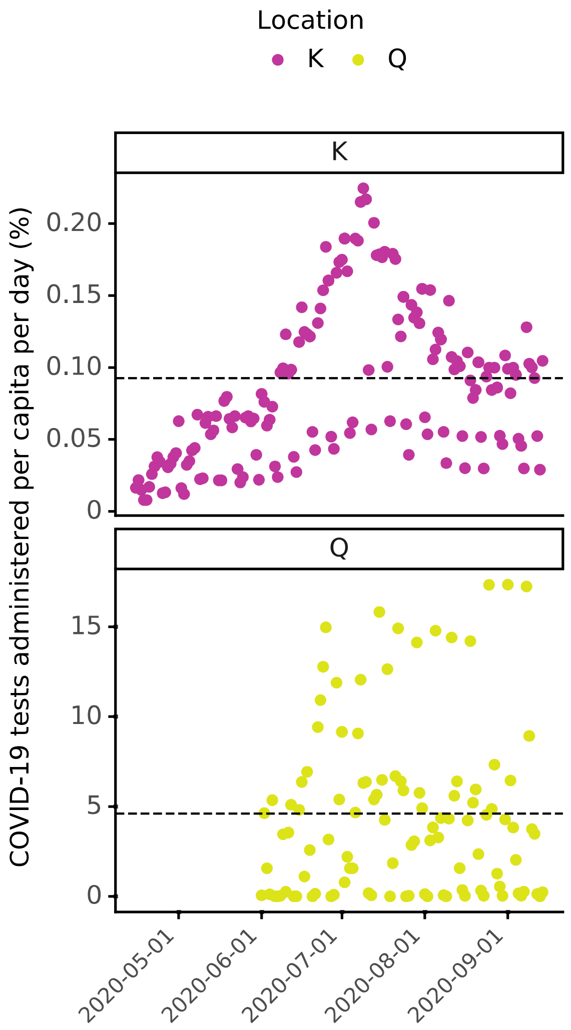


**Figure S15:** COVID-19 tests administered per capita (%) during the sampling period for two locations: K and Q, where dashed lines are plotted at the mean percentage of the population tested for the sampling period

### Author contributions

| Author | Contributions |
|---|---|
| Hannah D. Greenwald | Conceptualization; data analysis; conducted half of the qPCR labwork; manuscript writing; manuscript editing |
| Lauren C. Kennedy | Conceptualization; data analysis; conducted half of the qPCR labwork; manuscript writing; manuscript editing |
| Adrian Hinkle | Conducted sample extractions; manuscript editing |
| Vinson B. Fan | Contributed to assay development (18S rRNA assay, optimization of qPCR) |
| Oscar N. Whitney | Contributed to assay development (18S rRNA and crAssphage assays, optimization of extractions); manuscript editing |
| Sasha Harris-Lovett | Sample and data collection coordination with utilities, public health departments, and researchers; manuscript editing |
| Alexander Crits-Christoph | Contributed to assay development (optimization of extractions); manuscript editing |
| Avi I. Flamholz | Contributed to assay development (optimization of qPCR); manuscript editing |
| Basem Al-Shayeb | Contributed to assay development (optimization of extractions) |
| Lauren D. Liao | Contributed to qPCR pipeline code development |
| Dan Frost | Coordinated wastewater sample and physicochemical data collection |
| Daniel Brown | Coordinated clinical data collection |
| Chris Lynch | Coordinated clinical data collection |
| Jason Dow | Coordinated wastewater sample and physicochemical data collection |
| Mark Koekemoer | Coordinated wastewater sample and physicochemical data collection |
| Eileen White | Coordinated wastewater sample and physicochemical data collection |
| Alicia R. Chakrabati | Coordinated wastewater sample and physicochemical data collection |
| Matt Beyers | Coordinated clinical data collection |
| Payal Sarkar | Coordinated wastewater sample and physicochemical data collection |
| Rose Kantor | Conceptualization; data analysis; manuscript writing; manuscript editing |
| Kara L. Nelson | Conceptualization; data analysis; manuscript writing; manuscript editing |
